# The presence of SARS-CoV-2 RNA in different freshwater environments in urban settings determined by RT-qPCR: implications for water safety

**DOI:** 10.1101/2021.03.19.21253987

**Authors:** Jürgen Mahlknecht, Diego Alonso-Padilla, Edrick Ramos, Luisa Ma. Reyes, Mario Moises Álvarez

**Affiliations:** Centro del Agua para América Latina y el Caribe, Escuela de Ingeniería y Ciencias, Tecnologico de Monterrey, Monterrey, Nuevo Leon CP, 64849, Mexico; Centro de Biotecnología-FEMSA, Escuela de Ingeniería y Ciencias, Tecnologico de Monterrey, Monterrey, Nuevo Leon CP, 64849, Mexico

**Author notes:** **Corresponding authors:** Jurgen Mahlknecht, Mario M. Álvarez.

**Keywords:** SARS-CoV-2, freshwater, RT-qPCR, sucralose, Escherichia coli, water safety

## Abstract

This study is the first focused on the presence of SARS-CoV-2 in different freshwater environments in an urban setting. Groundwater and surface water reservoirs for drinking water as well as water from receiving rivers of the Monterrey Metropolitan Area were sampled repeatedly during a SARS-CoV-2 peak phase between October 2020 and January 2021, and viral RNA was measured by quantitative reverse transcription polymerase chain reaction. Forty-four percent of the groundwater samples had detectable viral loads between 2.6 and 38.3 copies/ml. A significant correlation between viral load and sucralose concentration in groundwater reaffirmed the hypothesis of leaching and infiltrating effluent from surface and/or failing sewage pipes and emphasized the importance of water disinfection. Twelve percent of the surface water dam samples tested positive for viral RNA, with values varying between 3.3 and 3.8 copies/ml. Finally, 13% of the river samples were positive for viral RNA, with concentrations ranging from 2.5 to 7.0 copies/ml. Untreated wastewater samples taken in the same period showed viral loads of up to 3535 copies/ml, demonstrating a dilution effect and/or wastewater facilities efficiency of three orders of magnitude. Variations in the viral loads in the groundwater and surface water over time and at the submetropolitan level generally reflected the reported trends in infection cases for Monterrey. The viral loads in the freshwater environments of Monterrey represent a low risk for recreational activities according to a preliminary risk assessment model. However, this result should not be taken lightly due to uncertainty regarding data and model constraints and the possibility of situations where the infection risk may increase considerably.

## 1. Introduction

Since the outbreak of the COVID-19 disease, various routes of transmission of SARS-CoV-2 have been verified and others have been hypothesized. Currently, the main transmission is known to occur between people through respiratory droplets (diameter >5-10 µm) produced by infected individuals when coughing or sneezing. Another presumed way of transmission is indirect contact with surfaces or objects in the immediate environment used by the infected person or on the infected person (Chan et al. 2020; Li et al. 2020; WHO, 2020).

An increasing number of studies have detected the presence of viral RNA in stool from COVID-19 patients (Wang et al., 2020; Kang et al., 2020; Xiao et al., 2020). Based on stool samples, Wu et al. (2020a) suggested that SARS-CoV-2 may replicate for 11 days in the gastrointestinal tract of patients even after samples from the respiratory tract become negative. According to another experiment, SARS-CoV-2 remained viable for 2 to 6 h in adult feces and up to 2 days in children’s feces (Liu 2020). This opens potential modes of fecal transmission.

Regarding the presence and persistence of SARS-CoV-2 in wastewater, there is sufficient evidence that indicates that wastewaters may contain both RNA fragments and viable particles of SARS-CoV-2 (Langone et al. 2020). Several studies have reported the new coronavirus in untreated and treated wastewater in the USA (Wu et al., 2020b; Nemudryi et al., 2020; Sherchan et al., 2020; Green et al., 2020; Peccia et al., 2020), Japan (Haramoto et al., 2020; Hata et al., 2020), France (Wurtzer et al., 2020; Torttier et al., 2020), Italy (La Rosa et al., 2020a; Rimoldi et al., 2020), Spain (Randazzo et al., 2020; Balboa et al., 2020), India (Kumar et al., 2020; Chakraborty et al., 2021), Pakistan (Sharif et al., 2020; Yaqub et al., 2020), Netherlands (Medema et al., 2020), Australia (Ahmed et al., 2020a), Turkey (Kocamemi et al., 2020), Israel (Bar-Or et al., 2020), Germany (Westhaus et al., 2020), and Czech Republic (Mlejnkova et al., 2020). Wastewater treatment plants (WWTPs) with tertiary disinfection have been found to be negative for SARS-CoV-2 (Rimoldi et al., 2020), while effluents from secondary treatments have been found to be positive (Randazzo et al., 2020; Rimoldi et al., 2020). The presence of SARS-CoV-2 RNA in sewage sludge was reported in a 10-week monitoring study in New Haven, Connecticut, USA (Peccia et al., 2020).

Although several authors have hypothesized about potential routes in water environments, to date, there exists little evidence of the presence of SARS-CoV-2 virus in freshwater (La Rosa et al. 2020b; Langone et al., 2020; Kumar et al., 2021). Rimoldi et al. (2020) detected viral RNA in three receiving rivers in the Milan area indicating the partial efficiency of the sewage system in the metropolitan area. Haramoto et al. (2020) collected three river samples between March and May 2020 in Japan and reported that no samples tested positive for SARS-CoV-2 RNA. Guerrero-Latorre et al. (2020) reported viral loads during a peak of the outbreak from three different sites of a river receiving untreated sewage from Quito, Ecuador. To our knowledge, to date, no evidence of the presence of the virus in surface water reservoirs and aquifers has been reported.

Water safety starts with the protection of water resources in catchment; therefore, it is mandatory to prevent surface and groundwater from coming into contact with fecal material. It is hypothesized that pathogen removal occurs in groundwater due to soil filtration, adsorption on sediment grains and progressive inactivation, and viruses in surface waters are exposed to several potentially inactivating stressors, including sunlight, oxidants, and predation by microorganisms (Langone et al., 2021). An ongoing research question is how persistent SARS-CoV-2 virus is in different water matrixes.

Chemical markers are indicators that may help evaluate the proper functioning of WWTPs and determine the level of human wastewater effluent in groundwater systems. The characteristics of an ideal wastewater indicator include: (i) source specificity, (ii) sustained effluent release because the indicator is not rapidly degraded by biological treatment processes, (iii) a demonstrated analytical methodology, (iv) no attenuation during transport, and (v) virtually zero background with a sufficiently large discharge to detection level ratio able to exceed receiving water dilution factors (Gasser et al., 2010; Oppenheimer et al., 2011). Several anthropogenic organic compounds with known characteristics have been used as chemical markers of pollutant loading due to their behavior as persistent aqueous organic pollutants (Benotti et al., 2009; Buerge et al., 2009). Among them, sucralose is one of the most popular artificial sweeteners and thus serves as a tracer of human wastewater, and its concentration is correlated with people connected to the sewage system (Kokotou et al., 2012; Voss et al., 2019). This organic compound is stable over a broad pH range and is heat stable, nonvolatile, highly polar and chiral. It is also strongly recalcitrant, degrading only under strongly oxidizing conditions, and is not metabolized by animals or microbes (Soh et al. 2011). These characteristics makes sucralose an excellent marker for human wastewater effluents, and it may help to confirm the presence of human pathogens such as SARS-CoV-2.

In the present study, we evaluated the presence of genetic material from SARS-CoV-2 RNA in different freshwater environments in the Monterrey Metropolitan Area (5.3 million inhabitants) in northern Mexico. The aim of the study was to perform a survey of viral dispersion and potential implications for the environment and public health during a peak phase of the epidemic. To address this goal, we collected untreated groundwater, river water and water from dams repeatedly between October 2020 and January 2021 and measured SARS-CoV-2 RNA by quantitative reverse transcription-polymerase chain reaction (qRT-PCR). For the groundwater, the concentration of the artificial sweetener sucralose was measured in parallel.

## 2. Materials and Methods

### 2.1 Study area

The Monterrey Metropolitan Area (MMA) is the second most important city in Mexico in terms of population and the economy. It comprises 12 municipalities with a total population of approximately 5.3 million inhabitants (INEGI, 2021). The climate is semiarid with a mean annual temperature and rainfall of 22.3°C and 622 mm, respectively, with a dry season (November-April) and rainy season (May-October). The urban area is bordered to the west and south by mountain ranges varying in composition from clastic marine to carbonate sedimentary rocks reaching elevations up to 2100 m above sea level (masl) (**Fig. 1**).

**Figure 1:**
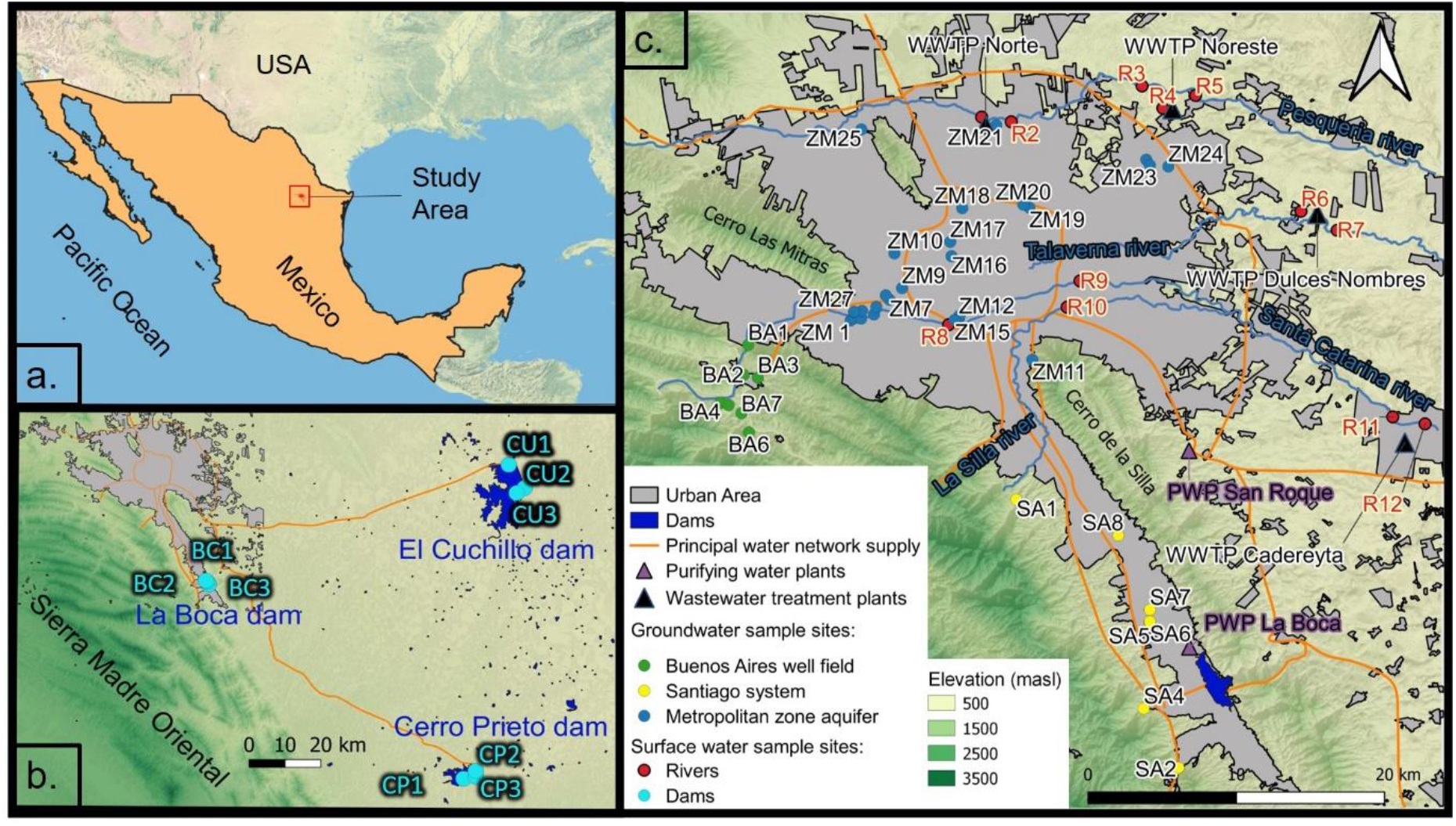
(a) Location of the study area; (b) regional view showing surface-water reservoirs with sampling points El Cuchillo (CU), Cerro Prieto (CP) and La Boca (BC) and (c) the urban area with the main features and sampling points of the groundwater systems Buenos Aires (BA), Santiago (SA) and Metropolitan zone (ZM), and urban rivers (R).

The MMA sits in a valley at 580 masl on Quaternary alluvial deposits eroded from the surrounding mountain ranges. The valley is mostly composed of fluvial and alluvial sedimentary deposits as terraces that occurred during accumulation-erosion cycles in the early Quaternary (Martinez and Werner, 1997).

Water for the MMA is supplied from surface water (58%) and groundwater (42%) reservoirs (SADM, 2021). Surface water is extracted from the El Cuchillo dam (4.69 m^3^/s), Cerro Prieto dam (2.83 m^3^/s), and La Boca dam (0.45 m^3^/s). Water from the El Cuchillo dam and Cerro Prieto dam is conveyed 108 km and 133 km to the MMA, respectively, while the La Boca dam connects to the Cerro Prieto aqueduct (**Fig. 1b**). Raw water from all three dams is purified before distribution throughout the city through two water-supply pipelines over 70 km in length each (**Fig. 1c**).

Groundwater is extracted from several aquifer units and wellfields and disinfected locally before being introduced into the supply network (Torres-Martinez et al. 2020) (**Fig. 1a**): The Buenos Aires (BA) well field (2.11 m^3^/s) located in a side valley close to the city consists of La Huasteca horizontal filtrating gallery and 23 deep wells with water table depths between 20 and 120 m below the ground, extracting water from Early Cretaceous limestone formations; the Santiago (SA) groundwater system (1.27 m^3^/s) consists of La Estanzuela spring and three horizontal filtrating galleries; the Monterrey Metropolitan Zone (ZM) aquifer (1.08 m^3^/s) includes wells throughout the metropolitan area, providing water from an unconfined aquifer which consists of altered lutite, conglomerate, gravel, sand and clay, with an average depth to groundwater of 20 m; finally, the Mina well field (1.20 m^3^/s) is located approximately 35 km northwest of the MMA.

Used domestic water is over 90% treated by public wastewater facilities that include primary and secondary stages in the treatment process. The most important wastewater treatment plants (WWTPs) are Dulces Nombres (7.5 m3/s), Norte (4.0 m3/s), Noreste (1.9 m3/s) and Cadereyta (0.25 m3/s) (**Fig. 1c**). All the noted WWTPs discharge the treated water directly or indirectly to the Pesquería River, except for the Cadereyta WWTP that discharges treated wastewater into the Santa Catarina River. Both rivers are tributaries of the San Juan River which in turn flows into the Rio Bravo/Rio Grande. The Pesquería and Santa Catarina Rivers had discharges that decreased from 5.5 to 4.4 m^3^/s at the Pesquería hydrometric station and from 5.6 to 2.5 m^3^/s at the Cadereyta hydrometric station between October 2020 and December 2020, respectively (SMN, 2021).

### 2.2 Field methods

Groundwater and surface water grab samples were collected at different sites and on different dates between October 2020 and January 2021. For the groundwater, 42 sites corresponding to production wells of supplying aquifer units of Monterrey (BA well field, ST system and ZM aquifer) were sampled initially between October 29 and November 3, 2020. Of these wells, 37 wells were public drinking water supply wells and five wells were used for industrial purposes. A subset of wells (n=10) was resampled two more times in cycles of approximately one month to observe changes over time (**Table 1**).

**Table 1:** Summary of sampling campaigns

| Freshwater type | Environments | Samples (first campaign) | Samples (second campaign) |
| --- | --- | --- | --- |
| Groundwater | Buenos aires well field, Santiago system, Zona Metropolitana aquifer | 40 | 10 |
| Rivers | Pesquería River, Santa Catarina River, La Silla River | 12 | 12 |
| Surface water reservoirs | El Cuchillo dam, Cerro Prieto dam, La Boca dam | 7 | 9 |

Similarly, samples were obtained from three sites of three surface water reservoirs supplying Monterrey (El Cuchillo, Cerro Prieto and La Boca) on October 22-23, 2020, and sampling was repeated two more times. Finally, a total of 12 river water grab samples were taken along the three urban rivers Pesquería, Santa Catarina and La Silla on December 10-11,2020, and his process was repeated on January 5-6, 2021. The river sites were selected strategically upstream and downstream of WWTP discharge into the rivers. For reference, 24-h composite samples of influent of a Dulces Nombres WWTP were taken weekly during the same period.

All samples were collected in sterile 125 ml HDPE bottles, stored at 4°C and analyzed within 48 h. SARS-CoV-2 is highly stable at 4°C (Chin et al.; 2020). Groundwater samples were included for analysis of sucralose, using 125 mlHDPE bottles.

### 2.3 Laboratory methods

#### 2.3.1 RNA and DNA extraction – QIAamp® Viral RNA Mini

We followed standard procedures to extract and purify nucleic acids from the water samples. Briefly, after viral thermal inactivation (95°C; 5 min), a volume of 500 µl of the water sample was centrifuged for 10 minutes at 1500 G. Then, a volume of 140 µl of the supernatant was added to a mix containing 0.56 µl of Buffer AVL solution (Qiagen, USA) and 5.6 µl of carrier RNA-AVE solution (Qiagen, USA) in a 1.5 ml microcentrifuge tube. This mix was vortexed for 15 seconds, incubated at room temperature (15–25°C) for 10 min and briefly centrifuged to remove drops from the interior surface of the lid. A volume of 560 µl of ethanol (96–100%) was added to the sample, and mixed by pulse-vortexing for 15 s.

After mixing, the tube was briefly centrifuged to remove drops from the interior surface of the lid. Then, this solution (∼630 µl) was filtered through a QIAamp Mini column (Qiagen, USA) to retain the nucleic acids originally present in the sample. The retained material was repeatedly washed with different buffer solutions to elute contaminants and purify the nucleic acids. Then, the solution was loaded into the column contained in a 2 mL collection tube, the cap of the tube was closed, and the tube with the column was centrifuged at 6000 x g (8000 rpm) for 1 min.

After centrifugation, the QIAamp Mini column was placed into a clean 2 ml collection tube, and the filtrate was discarded. In the first raising step, 500 µl of 96% ethanol was loaded into the column contained in the 2 mL collection tube, the cap of the tube was closed, and the tube with the column was centrifuged at 6000 x g (8000 rpm) for 1 min. Following these two centrifugation stages, 500 µl of buffer AW1 (Qiagen, USA) was added to the QIAamp Mini column, the cap of the container tube was closed, and the tube with the column was centrifuged at 6000 x g (8000 rpm) for 1 min. As before, the QIAamp Mini column was placed into a clean 2 ml collection tube, and the filtrate was discarded. In a fourth centrifugation cycle, a QIAamp Mini column was added to 500 µl buffer AW2 (Qiagen, USA), the cap of the container tube was closed, and the tube with the column was centrifuged at high speed (20,000 x g; 14,000 rpm) for 3 min.

Then, the QIAamp Mini column was placed in a clean 1.5 ml microcentrifuge tube and the filtrate was discarded. In a fifth centrifugation cycle, 60 µl buffer AVE (Qiagen, USA) equilibrated to room temperature was added to the QIAamp Mini column, the cap of the container tube was closed, and the tube with the column was centrifuged at a high speed (6,000 x g; 8,000 rpm) for 1 min.

For the DNA extraction, 500 µl of the water sample was centrifuged for 5 min at 5000 G; 400 µl of the centrifuge supernatant were discarded. The remaining 100 µl was added to 20 µl of proteinase K solution and 80 µl of buffer ATL (Qiagen, USA), vortexed, and incubated at 56°C for at least 1 hour. The remainder of the extraction protocol was analogous to that previously described.

#### 2.3.2 RNA and DNA amplification

We amplified RNA segments of SARS-CoV-2 using two sets of primers (commonly referred to as N1 and N2) in each amplification reaction. Both of these primers were directed to sequences that encode the N protein of SARS-CoV-2. These primer sets have been recommended and extensively used for the diagnosis of COVID-19 in human samples (Gonzalez-Gonzalez et al., 2020; Nalla et al., 2020) and wastewater (Medema et al., 2020; Wu et al., 2020b; Nemudryi et al., 2020; Randazzo et al., 2020; Haramoto et al., 2020; Scherchan et al., 2020; Peccia et al., 2020).

Similarly, we used two sets of primers to amplify the LAC and LAM regions of the genome of *Escherichia coli* in the same reaction. *E. coli* is used as a biological indicator of the presence of fecal content in water (Bej et al. 1990; Mo et al., 2002; Reza et al. 2014). The sequences of both the forward and reverse primers used are shown in **Table S1**.

Quantitative amplification was conducted in a quantitative PCR thermal cycle (Rotor gene Q 5plex, Qiagen, Germany). For the amplification of SARS-CoV-2 RNA sequences, the amplification mix (final volume of 20 µl) consisted of 10 µl of 2X QuantiNova Syber Green RT-Master Mix, 0.2 µl of QN SYBR Green RT-Mix, 1 µl of 10x primer mix (0.5 µM final concentration), and 8.8 µl of RNA extract. For the amplification of DNA sequences of *E. coli*, the amplification mix (final volume of 20 µl) consisted in 10 µl of 2X QuantiNova Syber Green RT-Master Mix, 1 µl of 10x primer mix (0.5 µM final concentration), and 9.0 µl of DNA extract. The amplification cycle consisted of 10 min of reverse transcription at 50°C and 2 min of amplification activation at 95°C, followed by 40 iterative cycles of denaturation for 5 s at 95°C and combined annealing and extension for 10 s at 60°C.

A calibration curve was constructed to establish the conversion between CT values and equivalent gene copies per milliliter (copies ml). For this purpose, we used commercial synthetic genetic material that contained the complete N gene from SARS-CoV-2 (Integrated DNA Technologies, Iowa, USA). Samples containing different concentrations of synthetic nucleic acids of SARS-CoV-2 (in the range of 10 to 100,000 copies ml^-1^) were prepared by successive dilutions from stocks. This plasmid has been used before as a positive control in amplification assays of SARS-CoV-2 genetic material (González-Gonzalez et al., 2021). The estimated lower limit of detection was ∼1 copy of the N gene of SARS-CoV-2 per mL of water. The lowest positive value was 2.5 copies/ml.

#### 2.3.3 Sucralose quantification

Sucralose is used as an artificial sweetener and useful tracer to demonstrate the presence of human wastewater in groundwater (Kokotou et al., 2012; Voss et al., 2019). Sucralose presence was determined using high performance liquid chromatography and mass spectrometric detection (HPLC-MS/MS) after solid-phase extraction (SOE). Isotope-labeled internal standards and an external calibration in tap water were used for quantification. Details of the analytical method are given in **Table S2**. The analysis was performed at DVGW-Technologiezentrum Wasser, Karlsruhe, Germany.

### 2.4 Monitoring of COVID-19 cases in Monterrey Metropolitan Area

To obtain an indication of the sensitivity of the monitoring of the urban water cycle, a proxy for the period prevalence of COVID-19 in the MMA was created using the reported number of COVID-19 cases per day (CONACyT, 2021) and the normalized cumulative number of reported COVID-19 cases per day for 2020. Normalization was performed by dividing the cumulative number of reported cases by the population size.

## 3. Results

### 3.1 Reported cases

The number of reported COVID-19 cases in each of the 12 municipalities and MMA shows that the pandemic evolved at different rates in each of the municipalities as it spread during 2020 (**Fig. 2a**). The first infection was reported on March 10, and the number of cases remained relatively low until mid-May, when another increase occurred, and starting from June 10, the infection maintained a constant increase in the MMA, with the exception of November, when the number of cases dropped. Santiago and Monterrey municipalities reported the most cases, followed by Santa Catarina, Guadalupe and San Nicolas.

**Figure 2:**
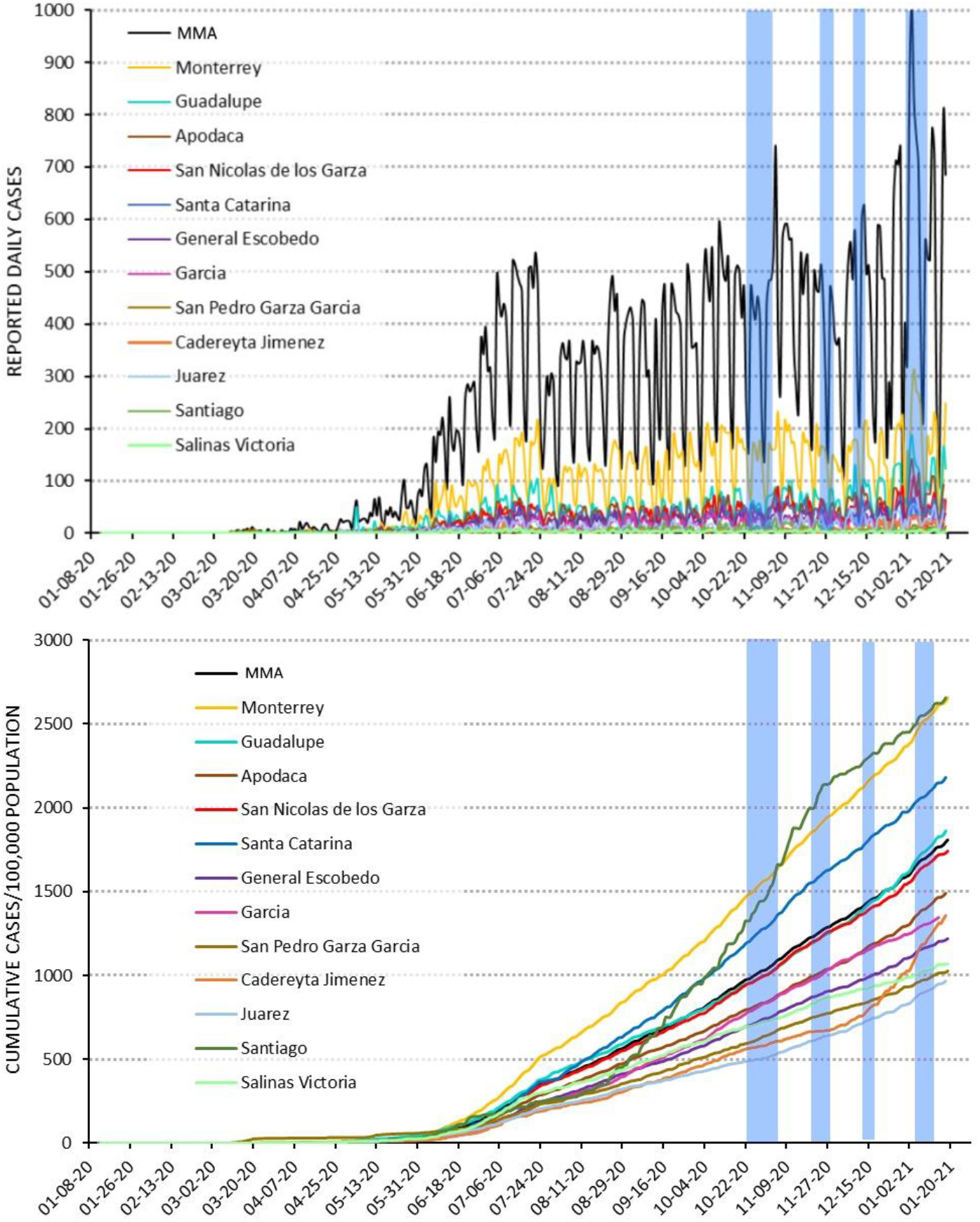
Reported cases for the MMA and its 12 municipalities: (a) reported daily cases of infection; and (b) normalized cumulative cases. Note: Date retrieved from CONACyT (2021). The vertical shaded blue lines indicate the sampling periods.

However, it is worth to noting that these numbers are not directly comparable to other countries or regions because the collection methods are not necessarily standardized, and the sampling efforts are probably different from and asynchronized respect to the real infection dates (Sims and Kasprzyk-Hordern, 2020). Freshwater sampling for this report was performed during the second peak of the outbreak of the epidemic: end of October, end of November 2020, mid-December 2020 and beginning January 2021 (**Fig. 2ab**).

### 3.2 Groundwater

Two field campaigns were performed for groundwater. Regarding the first campaign, the qRT–PCR concentration threshold (Ct) average values for SARS-CoV-2 ranged from 30.2 to over 40 (**Table 2**). Interestingly, nearly half of the samples (19 of 40) were positive, and 38% of the samples that tested positive had Ct values below the value of 33. In this study, a sample was arbitrarily defined as “positive” when a Ct value was detected in at least two of three replicates. Two of seven samples in the BA well field were detected positive, with Ct values of 30.2 and 32.4. Galeria 4 is a well at the entrance to the Huasteca highway, with a high urban development in the area prior to the entrance, while Pozo 39 is in the lower area of the Sierra Madre close to ranches and houses. Five out of eight samples in the SA system were reported to be positive, with a Ct value between 32.5 and 36.3. Estanzuela is in a woodland-rural environment, while Cola de Caballo Tunnels and San Francisco Tunnel represent horizontal galleries in piedmont shrubland. Finally, Margarita is a well located in an urban development area. Thirteen out of 26 production wells in the ZM aquifer had positive samples, with Ct values between 30.3 and 34.2. These sites are dispersed in the urbanized MMA. A trend showed a higher proportion of sites affected in the downstream area in the northeastern portion (Apodaca), and no positive samples in the southeastern portion (Contry) of the ZM aquifer.

**Table 2.** Summary of the results of determination of SARS-CoV-2 RNA and sucralose presence in groundwater in Monterrey. Note: The Ct value represents the average of s triplicate analysis for each sample, ‘n.d.’ indicates not detected and ‘-’ indicates not measured.

| Code | Site | Municipality | Geology | Land use | Groundwater level<br>(m below ground) | Campaign 1<br>29 Oct - 4 Nov 2020 |  | Campaign 2<br>26 - 30 Nov 2020 |
| --- | --- | --- | --- | --- | --- | --- | --- | --- |
|  |  |  |  |  |  | Ct<br>(cycles) | Sucralose<br>(µg/L) | Ct<br>(cycles) |
| BA1 | Galeria 4 | Santa Catarina | Limestone | Urban Area | 0.0 | 30.2 | n.d. | n.d. |
| BA2 | Pozo 39 | Santa Catarina | Limestone | Desert Shrubland | 43.0 | 32.3 | n.d. | n.d. |
| BA3 | Pozo 28 | Santa Catarina | Limestone | Desert Shrubland | 43.0 | n.d. | n.d. | - |
| BA4 | Pozo 1 | Santa Catarina | Alluvial Deposits | Piedmont Shrubland | 40.7 | n.d. | n.d. | - |
| BA5 | Pozo 4 | Santa Catarina | Alluvial Deposits | Piedmont Shrubland | 43.1 | n.d. | n.d. | - |
| BA6 | Pozo 14 | Santa Catarina | Alluvial Deposits | Desert Shrubland | 75.0 | n.d. | n.d. | - |
| BA7 | Pozo 2 | Santa Catarina | Alluvial Deposits | Desert Shrubland | 0.0 | n.d. | n.d. | - |
| SA1 | Estanzuela | Santiago | Shale | Urban Area | 0.0 | 32.5 | n.d. | - |
| SA2 | Tunel 1 Cola de Caballo | Santiago | Limestone | Piedmont Shrubland | 0.0 | 32.6 | n.d. | - |
| SA3 | Tunel 2 Cola de Caballo | Santiago | Shale | Mixed woodland | 0.0 | 33.9 | n.d. | - |
| SA4 | Tunel San Francisco | Santiago | Shale | Piedmont Shrubland | 0.0 | 32.8 | n.d. | - |
| SA5 | Andares | Santiago | Alluvial Deposits | Urban Area | 11.8 | n.d. | 0.07 | - |
| SA6 | Condado de Asturias | Santiago | Alluvial Deposits | Urban Area | 12.3 | n.d. | n.d. | - |
| SA7 | Pozo Rodriguez | Santiago | Alluvial Deposits | Urban Area | 15.9 | n.d. | n.d. | - |
| SA8 | Pozo Margaritas | Santiago | Alluvial Deposits | Urban Area | 11.3 | 36.3 | 2.90 | - |
| ZM1 | Auditorio San Pedro | San Pedro | Alluvial Deposits | Urban Area | 21.2 | 30.5 | 0.54 | n.d. |
| ZM2 | Humberto Lobo | San Pedro | Alluvial Deposits | Urban Area | 14.4 | n.d. | 1.80 | - |
| ZM3 | Suchiate II | San Pedro | Alluvial Deposits | Urban Area | 10.9 | n.d. | 0.51 | - |
| ZM4 | Pozo Profundo Monterrey I | Monterrey | Alluvial Deposits | Urban Area | 20.9 | 30.3 | n.d. | - |
| ZM5 | Pozo Profundo Monterrey II | Monterrey | Alluvial Deposits | Urban Area | 20.1 | 31.2 | n.d. | n.d. |
| ZM6 | San Jerónimo II | Monterrey | Alluvial Deposits | Urban Area | 26.1 | n.d. | 2.70 | - |
| ZM7 | Pozo Profundo Monterrey III | Monterrey | Alluvial Deposits | Urban Area | 17.4 | n.d. | n.d. | - |
| ZM8 | Pozo Profundo Monterrey VI | Monterrey | Shale | Urban Area | 9.8 | n.d. | n.d. | - |
| ZM9 | Hospital Civil Norte | Monterrey | Alluvial Deposits | Urban Area | 115.0 | n.d. | 1.20 | - |
| ZM10 | Lincoln II | Monterrey | Alluvial Deposits | Urban Area | 22.2 | 33.2 | 0.67 | 34.0 |
| ZM11 | Monterrey V | Guadalupe | Limestone | Urban Area | 69.1 | n.d. | n.d. | - |
| ZM12 | Metro Rey Oriente | Monterrey | Alluvial Deposits | Urban Area | 6.2 | n.d. | 0.44 | - |
| ZM13 | Metro Rey Poniente | Monterrey | Alluvial Deposits | Urban Area | 7.9 | n.d. | 0.46 | - |
| ZM14 | Macro Plaza II | Monterrey | Alluvial Deposits | Urban Area | 3.9 | 31.5 | 0.46 | n.d. |
| ZM15 | Plaza Hidalgo | Monterrey | Alluvial Deposits | Urban Area | 12.3 | 34.2 | 0.46 | - |
| ZM16 | Somero California II | San Nicolás | Alluvial Deposits | Urban Area | 14.8 | 30.8 | 1.00 | 33.1 |
| ZM17 | Estadio Beisbol | San Nicolás | Alluvial Deposits | Mixed woodland | 14.2 | n.d. | 0.43 | - |
| ZM18 | Somero El Roble | San Nicolás | Alluvial Deposits | Urban Area | 22.9 | n.d. | 0.49 | - |
| ZM19 | Somero Puentes Avenida | San Nicolás | Alluvial Deposits | Urban Area | 44.0 | n.d. | 1.40 | - |
| ZM20 | Somero Puentes II | San Nicolás | Alluvial Deposits | Urban Area | 11.2 | 31.5 | 1.30 | 33.1 |
| ZM21 | Tecno Centro I | San Nicolás | Conglomerate | Urban/Industrial | 10.3 | 30.7 | 0.77 | - |
| ZM22 | Papa 02 | Apodaca | Alluvial Deposits | Urban | 13.3 | 30.9 | 0.20 | n.d. |
| ZM23 | Papa 03 | Apodaca | Alluvial Deposits | Urban | 13.5 | 30.7 | 0.16 | n.d. |
| ZM24 | Pozo PIMSA II | Apodaca | Alluvial Deposits | Urban/Industrial | 7.4 | n.d. | 0.97 | - |
| ZM25 | Topo Chico III | Monterrey | Limestone | Urban Area | 25.8 | 33.0 | 0.10 | - |

Sucralose was detected in 22 out of 40 samples (55%) (**Table 2**), and its concentrations varied between 0.07 and 2.9 µg/l. In the BA well field, which represents desert and piedmont shrubland with a low population density, none of the samples had detectable levels. In the SA system, one site (Andares) had concentrations of sucralose close to the detection limit, and one site (Margaritas) had a sample with one of the highest concentrations. These sites represented residential areas. In the urbanized ZM aquifer, 20 out of 25 well sites (80%) had detectable concentrations of sucralose, whose values ranged between 0.1 and 2.7 µg/l. These results are generally consistent with the land use distribution, and all except one site in urbanized or industrial plots had samples with sucralose. In addition, we found a significant correlation between sucralose and Ct values (r2=0.62, n=0.043) but no correlation between Ct values and groundwater depth.

The samples that were positive in the first sampling campaign and not located close to each other were repeated for a second campaign (**Table 2**). In the second sampling campaign only 3 out of 10 sites tested positive for SARS-CoV-2. This suggests that groundwater was less affected on the second sampling date, and only three sites had samples that were consistently positive on both dates, namely, California 2, Lincoln 2 and Puentes 1 in Monterrey municipality. It is notable that the depth-to-water table of these sites was less than 22 m.

### 3.3 Surface water

Two sampling campaigns were performed in surface water reservoirs between the end of October and mid-December 2020 (**Table 3**). For the first period in October 2020 none of the samples were detected positive. For the second sampling period two sites had samples that tested positive, one in the La Boca dam (33.8) and another in the Cerro Prieto dam (33.6). It was not possible to analyze the correlation between the Ct values of SARS-CoV-2 and *E. coli* because only two pairs had quantitative data.

**Table 3.** Results of determination of SARS-CoV-2 RNA and E. coli presence in surface water reservoirs. Note: The Ct value represents the average of triplicate analysis for each sample, ‘n.d.’ indicates not detected, and ‘-’ indicates not measured.

| ID | Site | Campaign 1<br>22 - 23 Oct 2020 |  | Campaign 2<br>14 - 15 Dec 2020 |  |
| --- | --- | --- | --- | --- | --- |
|  |  | Ct (SARS-CoV-2) | Ct (*E. coli*) | Ct (SARS-CoV-2) | Ct (*E. coli*) |
|  |  | (cycles) | (cycles) | (cycles) | (cycles) |
| BO1 | La Boca 1 | n.d. | 31.3 | n.d. | 34.5 |
| BO2 | La Boca 2 | - | - | n.d. | 33.9 |
| BO3 | La Boca 3 | - | - | 33.8 | 32.4 |
| CP1 | Cerro Prieto 1 | n.d. | 29.7 | n.d. | 32.5 |
| CP2 | Cerro Prieto 2 | n.d. | 30.6 | n.d. | 32.2 |
| CP3 | Cerro Prieto 3 | n.d. | 30.7 | 33.6 | 33.2 |
| CU1 | El Cuchillo 1 | n.d. | 31.5 | n.d. | 31.5 |
| CU2 | El Cuchillo 2 | n.d. | 30.8 | n.d. | 31.4 |
| CU3 | El Cuchillo 3 | n.d. | 30.8 | n.d. | 33.0 |

With respect to river water, two sampling campaigns were performed in December 2020 and in January 2021. In December, three out of twelve samples tested positive, with Ct values ranging from 32.7 to 34.2. The sites with positive values were the Pesqueria River downstream of WWTP Norte, Santa Catarina River upstream of WWTP Cadereyta, and La Silla River upstream of Tolteca Park. For the second sampling period, two out of twelve samples were positive, namely, the Pesquería River upstream WWTP Norte and La Silla River at upstream of Tolteca Park (**Table 4**). The result for La Silla River was notable because this river receives no treated wastewaters of domestic origin. The Ct values of SARS-CoV-2 correlated with those of *E. coli* (r2=0.75, n=0.088); however the correlation was weak due to the low number of pairs.

**Table 4.** Results of the determination of SARS-CoV-2 RNA presence in rivers in the MMA. Note: The Ct value represents the average of triplicate analysis for each sample and ‘n.d.’ means not detected.

| ID | Site | Campaign 1<br>10 - 11 Dic 2020 |  | Campaign 2<br>5-6 Jan 2021 |  |
| --- | --- | --- | --- | --- | --- |
|  |  | Ct SARS-CoV-2<br>(cycles) | Ct E. Coli<br>(cycles) | Ct SARS-CoV-2<br>(cycles) | Ct E. Coli<br>(cycles) |
| R1 | Pesquería River upstream WWTP Norte | n.d. | 31.7 | 35.9 | 33.8 |
| R2 | Pesquería River downstream WWTP Norte | 34.2 | 28.9 | n.d. | 31.5 |
| R3 | Channel upstream WWTP Noreste | n.d. | 31.0 | n.d. | 31.0 |
| R4 | Pesquería River upstream WWTP Noreste | n.d. | 30.9 | n.d. | 30.9 |
| R5 | Pesquería River downstream WWTP Noreste | n.d. | 31.4 | n.d. | 31.4 |
| R6 | Channel upstream WWTP Dulces Nombres | n.d. | 29.5 | n.d. | 29.5 |
| R7 | Channel downstream WWTP Dulces Nombres | n.d. | 29.1 | n.d. | 29.1 |
| R8 | Santa Catarina River downtown | n.d. | 32.5 | n.d. | 32.5 |
| R9 | Santa Catarina River after downtown | n.d. | 33.1 | n.d. | 33.1 |
| R10 | La Silla River | 33.9 | 32.2 | 37.5 | 34.5 |
| R11 | Santa Catarina River upstream WWTP Cadereyta | 32.7 | 29.4 | n.d. | 31.1 |
| R12 | Santa Catarina River downstream WWTP Cadereyta | n.d. | 30.4 | n.d. | 30.4 |

### 3.4 Wastewater

For reference, untreated wastewater from the influent of the Dulces Nombres WWTP was measured for SARS-CoV-2. Between October 25, 2020, and December 13, 2020, 3 out of 8 samples (38%) were positive. The Ct value of positive samples ranged from 23.5 to 31.2.

## 4. Discussion

### 4.1 Contextualization of the findings in freshwater environments

This is the first study that quantifies the presence of SARS-CoV-2 in different freshwater environments of an urban setting. Previous studies that aimed to detect the virus in freshwater focused on receiving rivers (**Table 5**). For example, Rimoldi et al. (2020) collected grab samples at three sites of receptor rivers in the Milan area on April 14 and 22, 2020. In the first sampling round, all three samples were positive, while in the second round only one out of three samples was positive. A quantitative analysis was not performed. Similarly, Haramoto et al. (2020) collected grab water samples in a river in Yamanashi Prefecture, Japan, on three different occasions between April 22 and May 7, 2020; they reported that no sample tested positive for SARS-CoV-2 RNA.

**Table 5.**
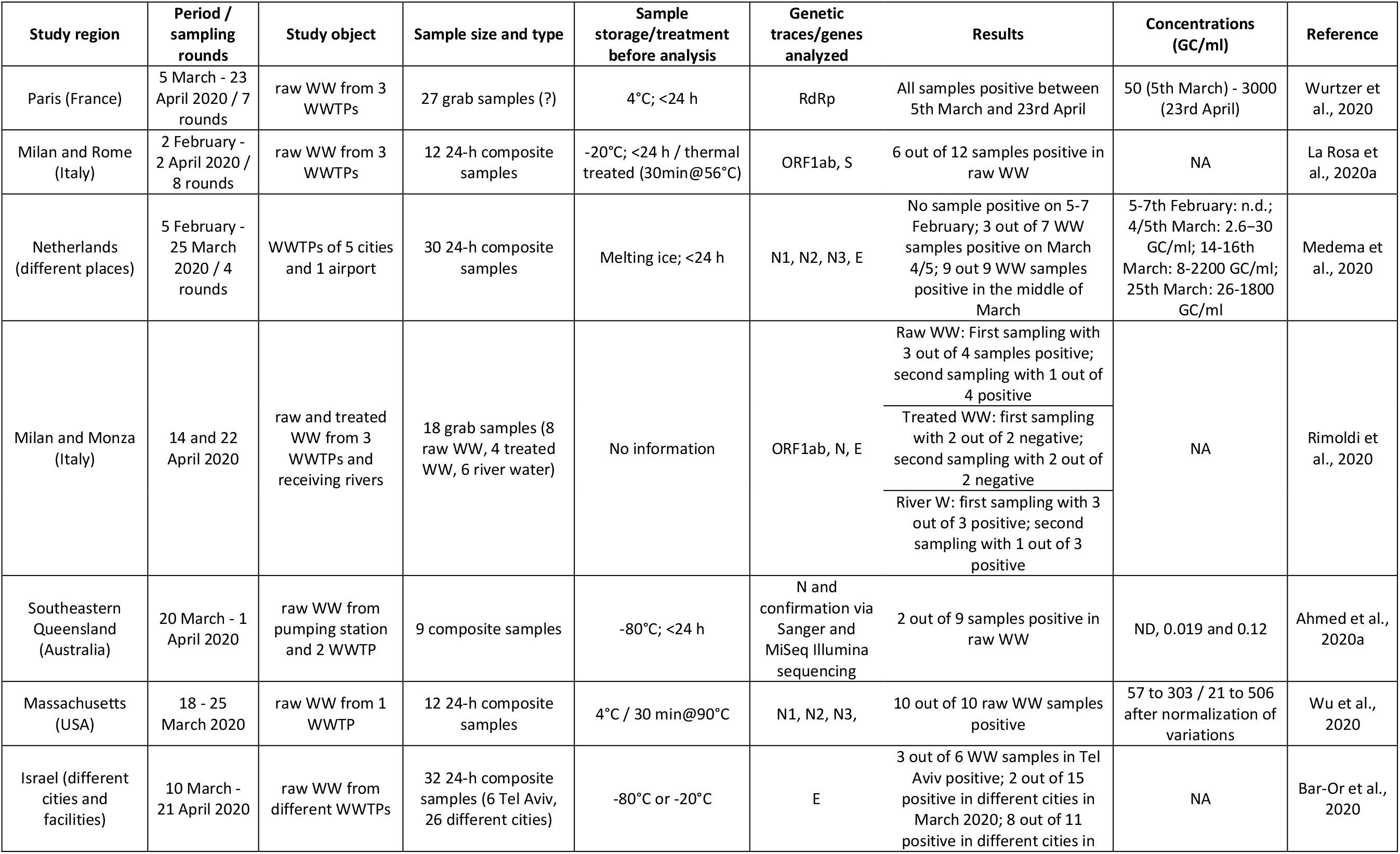
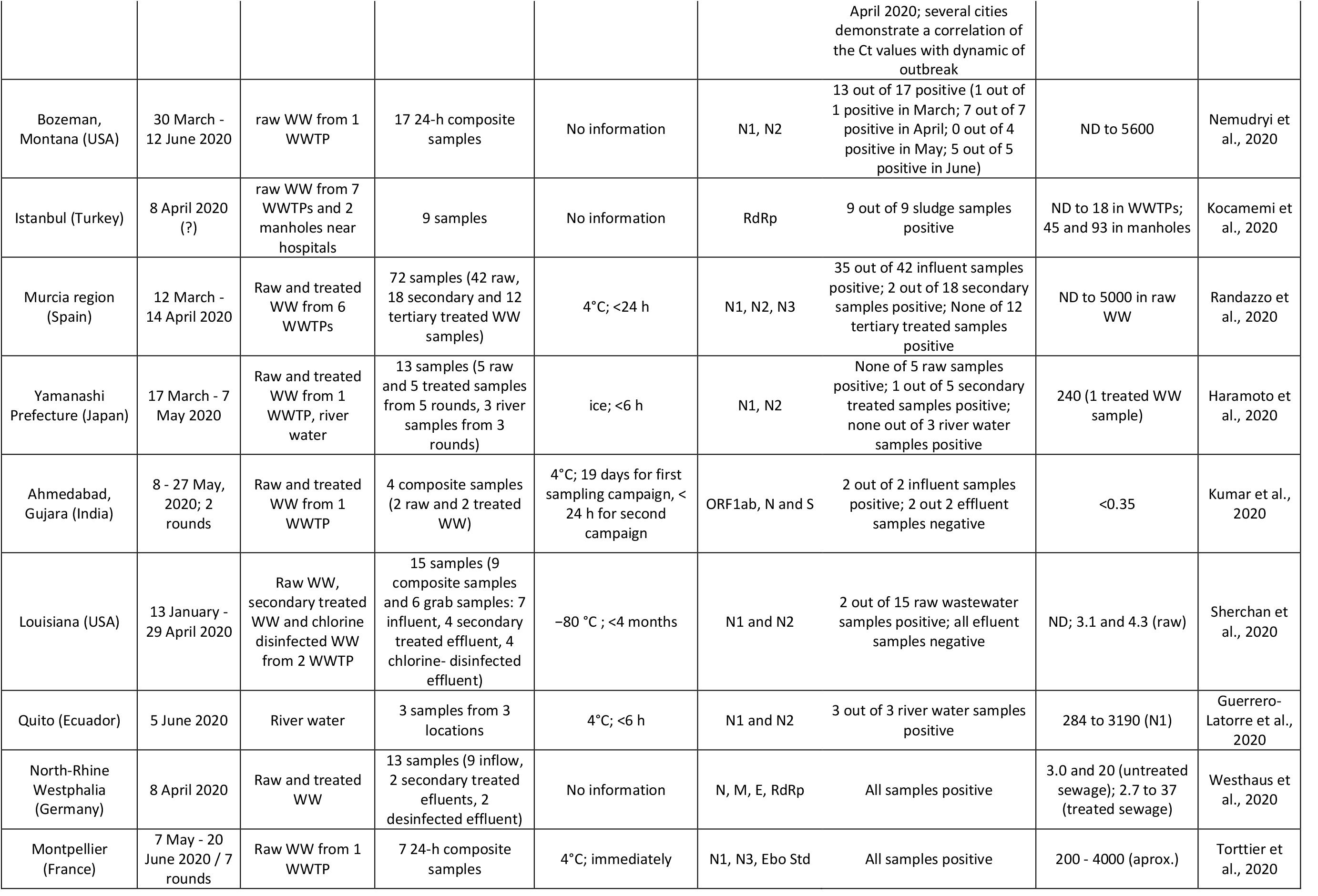
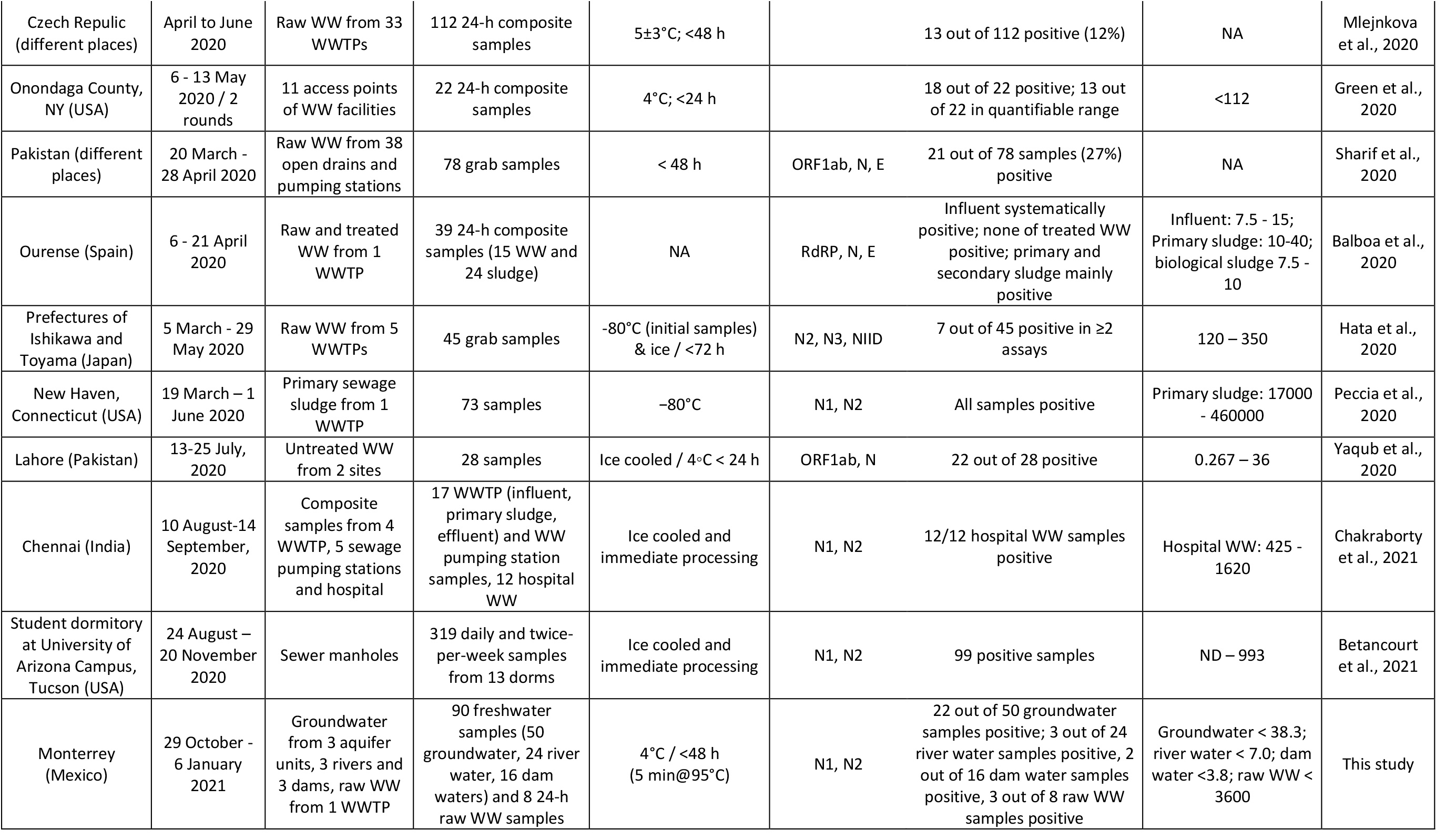
Selected studies on municipal wastewater/sludge and receiving river waters. Note: ‘NA’ means not applied and ‘WW’ indicates wastewater.

| Study region | Period / sampling rounds | Study object | Sample size and type | Sample storage/treatment before analysis | Genetic traces/genes analyzed | Results | Concentrations (GC/ml) | Reference |
| --- | --- | --- | --- | --- | --- | --- | --- | --- |
| Paris (France) | 5 March - 23 April 2020 / 7 rounds | raw WW from 3 WWTPs | 27 grab samples (?) | 4°C; <24 h | RdRp | All samples positive between 5th March and 23rd April | 50 (5th March) - 3000 (23rd April) | Wurtzer et al., 2020 |
| Milan and Rome (Italy) | 2 February - 2 April 2020 / 8 rounds | raw WW from 3 WWTPs | 12 24-h composite samples | -20°C; <24 h / thermal treated (30min@56°C) | ORF1ab, S | 6 out of 12 samples positive in raw WW | NA | La Rosa et al., 2020a |
| Netherlands (different places) | 5 February - 25 March 2020 / 4 rounds | WWTPs of 5 cities and 1 airport | 30 24-h composite samples | Melting ice; <24 h | N1, N2, N3, E | No sample positive on 5-7 February; 3 out of 7 WW samples positive on March 4/5; 9 out 9 WW samples positive in the middle of March | 5-7th February: n.d.; 4/5th March: 2.6–30 GC/ml; 14-16th March: 8-2200 GC/ml; 25th March: 26-1800 GC/ml | Medema et al., 2020 |
| Milan and Monza (Italy) | 14 and 22 April 2020 | raw and treated WW from 3 WWTPs and receiving rivers | 18 grab samples (8 raw WW, 4 treated WW, 6 river water) | No information | ORF1ab, N, E | Raw WW: First sampling with 3 out of 4 samples positive; second sampling with 1 out of 4 positive<br>Treated WW: first sampling with 2 out of 2 negative; second sampling with 2 out of 2 negative<br>River W: first sampling with 3 out of 3 positive; second sampling with 1 out of 3 positive | NA | Rimoldi et al., 2020 |
| Southeastern Queensland (Australia) | 20 March - 1 April 2020 | raw WW from pumping station and 2 WWTP | 9 composite samples | -80°C; <24 h | N and confirmation via Sanger and MiSeq Illumina sequencing | 2 out of 9 samples positive in raw WW | ND, 0.019 and 0.12 | Ahmed et al., 2020a |
| Massachusetts (USA) | 18 - 25 March 2020 | raw WW from 1 WWTP | 12 24-h composite samples | 4°C / 30 min@90°C | N1, N2, N3, | 10 out of 10 raw WW samples positive | 57 to 303 / 21 to 506 after normalization of variations | Wu et al., 2020 |
| Israel (different cities and facilities) | 10 March - 21 April 2020 | raw WW from different WWTPs | 32 24-h composite samples (6 Tel Aviv, 26 different cities) | -80°C or -20°C | E | 3 out of 6 WW samples in Tel Aviv positive; 2 out of 15 positive in different cities in March 2020; 8 out of 11 positive in different cities in | NA | Bar-Or et al., 2020 |
|  |  |  |  |  |  | April 2020; several cities demonstrate a correlation of the Ct values with dynamic of outbreak |  |  |
| Bozeman, Montana (USA) | 30 March - 12 June 2020 | raw WW from 1 WWTP | 17 24-h composite samples | No information | N1, N2 | 13 out of 17 positive (1 out of 1 positive in March; 7 out of 7 positive in April; 0 out of 4 positive in May; 5 out of 5 positive in June) | ND to 5600 | Nemudryi et al., 2020 |
| Istanbul (Turkey) | 8 April 2020 (?) | raw WW from 7 WWTPs and 2 manholes near hospitals | 9 samples | No information | RdRp | 9 out of 9 sludge samples positive | ND to 18 in WWTPs; 45 and 93 in manholes | Kocamemi et al., 2020 |
| Murcia region (Spain) | 12 March - 14 April 2020 | Raw and treated WW from 6 WWTPs | 72 samples (42 raw, 18 secondary and 12 tertiary treated WW samples) | 4°C; <24 h | N1, N2, N3 | 35 out of 42 influent samples positive; 2 out of 18 secondary samples positive; None of 12 tertiary treated samples positive | ND to 5000 in raw WW | Randazzo et al., 2020 |
| Yamanashi Prefecture (Japan) | 17 March - 7 May 2020 | Raw and treated WW from 1 WWTP, river water | 13 samples (5 raw and 5 treated samples from 5 rounds, 3 river samples from 3 rounds) | ice; <6 h | N1, N2 | None of 5 raw samples positive; 1 out of 5 secondary treated samples positive; none out of 3 river water samples positive | 240 (1 treated WW sample) | Haramoto et al., 2020 |
| Ahmedabad, Gujara (India) | 8 - 27 May, 2020; 2 rounds | Raw and treated WW from 1 WWTP | 4 composite samples (2 raw and 2 treated WW) | 4°C; 19 days for first sampling campaign, < 24 h for second campaign | ORF1ab, N and S | 2 out of 2 influent samples positive; 2 out of 2 effluent samples negative | <0.35 | Kumar et al., 2020 |
| Louisiana (USA) | 13 January - 29 April 2020 | Raw WW, secondary treated WW and chlorine disinfected WW from 2 WWTP | 15 samples (9 composite samples and 6 grab samples: 7 influent, 4 secondary treated effluent, 4 chlorine- disinfected effluent) | -80 °C ; <4 months | N1 and N2 | 2 out of 15 raw wastewater samples positive; all effluent samples negative | ND; 3.1 and 4.3 (raw) | Sherchan et al., 2020 |
| Quito (Ecuador) | 5 June 2020 | River water | 3 samples from 3 locations | 4°C; <6 h | N1 and N2 | 3 out of 3 river water samples positive | 284 to 3190 (N1) | Guerrero-Latorre et al., 2020 |
| North-Rhine Westphalia (Germany) | 8 April 2020 | Raw and treated WW | 13 samples (9 inflow, 2 secondary treated effluents, 2 disinfected effluent) | No information | N, M, E, RdRp | All samples positive | 3.0 and 20 (untreated sewage); 2.7 to 37 (treated sewage) | Westhaus et al., 2020 |
| Montpellier (France) | 7 May - 20 June 2020 / 7 rounds | Raw WW from 1 WWTP | 7 24-h composite samples | 4°C; immediately | N1, N3, Ebo Std | All samples positive | 200 - 4000 (aprox.) | Torttier et al., 2020 |
| Czech Republic (different places) | April to June 2020 | Raw WW from 33 WWTPs | 112 24-h composite samples | 5±3°C; <48 h |  | 13 out of 112 positive (12%) | NA | Mlejnkova et al., 2020 |
| Onondaga County, NY (USA) | 6 - 13 May 2020 / 2 rounds | 11 access points of WW facilities | 22 24-h composite samples | 4°C; <24 h |  | 18 out of 22 positive; 13 out of 22 in quantifiable range | <112 | Green et al., 2020 |
| Pakistan (different places) | 20 March - 28 April 2020 | Raw WW from 38 open drains and pumping stations | 78 grab samples | < 48 h | ORF1ab, N, E | 21 out of 78 samples (27%) positive | NA | Sharif et al., 2020 |
| Ourense (Spain) | 6 - 21 April 2020 | Raw and treated WW from 1 WWTP | 39 24-h composite samples (15 WW and 24 sludge) | NA | RdRP, N, E | Influent systematically positive; none of treated WW positive; primary and secondary sludge mainly positive | Influent: 7.5 - 15; Primary sludge: 10-40; biological sludge 7.5 - 10 | Balboa et al., 2020 |
| Prefectures of Ishikawa and Toyama (Japan) | 5 March - 29 May 2020 | Raw WW from 5 WWTPs | 45 grab samples | -80°C (initial samples) & ice / <72 h | N2, N3, NIID | 7 out of 45 positive in ≥2 assays | 120 – 350 | Hata et al., 2020 |
| New Haven, Connecticut (USA) | 19 March – 1 June 2020 | Primary sewage sludge from 1 WWTP | 73 samples | -80°C | N1, N2 | All samples positive | Primary sludge: 17000 - 460000 | Peccia et al., 2020 |
| Lahore (Pakistan) | 13-25 July, 2020 | Untreated WW from 2 sites | 28 samples | Ice cooled / 4°C < 24 h | ORF1ab, N | 22 out of 28 positive | 0.267 – 36 | Yaqub et al., 2020 |
| Chennai (India) | 10 August-14 September, 2020 | Composite samples from 4 WWTP, 5 sewage pumping stations and hospital | 17 WWTP (influent, primary sludge, effluent) and WW pumping station samples, 12 hospital WW | Ice cooled and immediate processing | N1, N2 | 12/12 hospital WW samples positive | Hospital WW: 425 - 1620 | Chakraborty et al., 2021 |
| Student dormitory at University of Arizona Campus, Tucson (USA) | 24 August – 20 November 2020 | Sewer manholes | 319 daily and twice-per-week samples from 13 dorms | Ice cooled and immediate processing | N1, N2 | 99 positive samples | ND – 993 | Betancourt et al., 2021 |
| Monterrey (Mexico) | 29 October - 6 January 2021 | Groundwater from 3 aquifer units, 3 rivers and 3 dams, raw WW from 1 WWTP | 90 freshwater samples (50 groundwater, 24 river water, 16 dam waters) and 8 24-h raw WW samples | 4°C / <48 h (5 min@95°C) | N1, N2 | 22 out of 50 groundwater samples positive; 3 out of 24 river water samples positive, 2 out of 16 dam water samples positive, 3 out of 8 raw WW samples positive | Groundwater < 38.3; river water < 7.0; dam water < 3.8; raw WW < 3600 | This study |

Guerrero-Latorre et al. (2020) reported viral loads during a peak of the outbreak (June 5, 2020) from three different sites of a river receiving untreated sewage from Quito city. The authors used RT-qPCR for these determinations and two different primer sets, namely N1 and N2. All samples were found to be positive, and the values ranged from 284 to 3190 GC/ml and from 207 to 2230 GC/ml in assays using the N1 and N2 target regions, respectively. These values could be related clearly to COVID-19 cases reported in the contributing areas.

### 4.2 Explanation of viral loads in receiving waters

In the present study, 13% of all river water samples (3 out of 24) were positive regarding viral RNA, and the viral RNA amounts in the positive samples varied between 2.5 and 7.0 GC/ml (Fig. 3ab). Importantly, during this period no significant rainfall was recorded in the Monterrey area that could have had an impact on virus concentration in the water. These loads are two to three orders of magnitude lower than those reported by Guerrero-Latorre et al. (2020) for Quito’s river. This could be because Monterrey treats more than 95% of its municipal wastewater, while the urban rivers of Quito are impacted by the direct discharge of sewage water from the city (3 million inhabitants). Similarly, the negative results derived from the analysis of river water samples from Yamanashi Prefecture, Japan (Haramoto et al. 2020) and Milan, Italy (Rimoldi et al. 2020) could be attributed to the fact that both studies collected water from rivers receiving treated wastewater.

**Figure 3:**
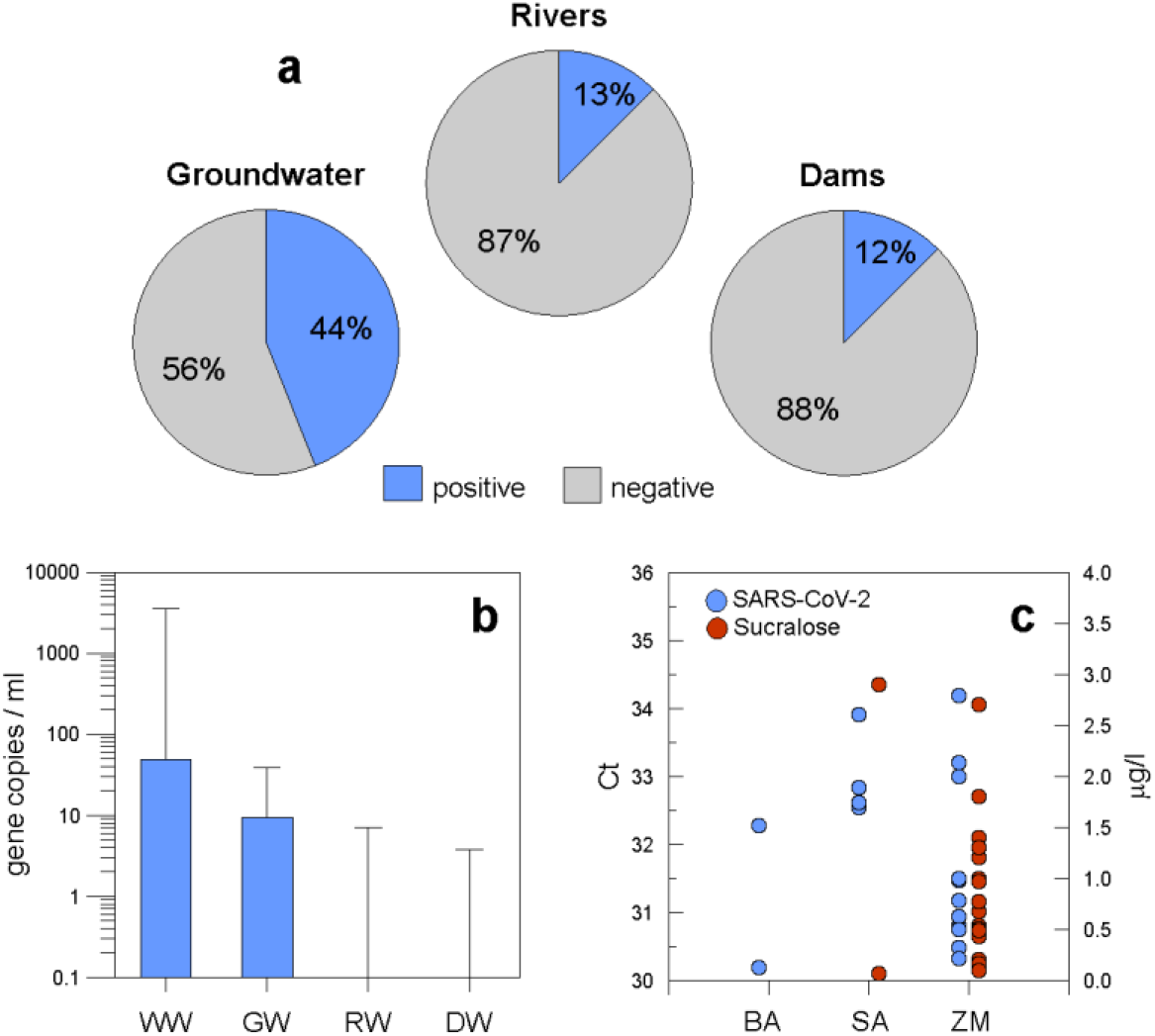
(a) Share of positive and negative samples in the different freshwater environments; (b) Boxplot of viral loads of different water/wastewater types (WW=wastewater, GW=groundwater, RW=river water; DW=dam water); (c) SARS-CoV2 and sucralose scatter graph (BA=Buenos Aires, SA=Santiago; ZM=Zona Metropolitana).

It is expected that wastewater from WWTPs that is completely treated would test negative. Thus, the occurrence of SARS-CoV-2 RNA in a few samples in La Silla and Pesquería River water could stem from different wastewater sources coexisting in the same basin. For example, aliquots of untreated sewage can be present because of illegal discharges, local malfunctions of sewerage systems, and their increased relative contribution during dry periods (Mosley et al. 2015). The lack of separation of urban runoff water from the domestic effluents, which causes combined sewer overflows (CSOs), could also be a reason for this occurrence of viral loads (Rimoldi et al. 2020). CSOs occur usually during high rainfall events. However, the accumulated rainfall between December 2020 and January 2021 in Monterrey was only 3 mm.

Another reason for the high aliquots of untreated sewage in river water could be the organization of local football derbies, whose high loads in short time periods may overburden the capacity of WWTPs to release untreated wastewater to the Pesquería river (SADM, 2020). The case of the La Silla River is notable because it receives no relevant treated municipal wastewater due to sanitary drainage to the other two rivers; therefore illegal discharges or a local sewage system malfunction is a plausible explanation for the presence of SARS-CoV-2 genetic material in this water course.

Regarding dam water, only 12% of the samples (2 out of 16) tested positive for SARS-CoV-2 RNA (Fig. 3a), with no positive result in the first campaign (22-23 October 2020). The positive samples (which contained 3.3 and 3.8 viral copies/ml) occurred during the second campaign (14-15 December 2020) and only at one site in the La Boca and at one site in the Cerro Prieto dam, respectively. In both cases, a village is located nearby, which suggests that the presence of the virus might be due to failure of the local sewage system. The observed values were comparable to the range of values in the urban rivers in Monterrey. The lack of viral loads in the first campaign and the presence of viral loads at two of the nine sites in the second campaign may reflect the increasing trend in reported cases of infection in the corresponding municipalities during the same period (**Fig. 1a**).

### 4.3 Viral load in groundwater reaffirms human sewage impact

The number of groundwater samples containing detectable SARS-CoV-2 RNA was surprisingly high. Twenty-two out of 50 samples (44%) had viral loads between 2.9 and 38.3 GC/ml (**Fig. 3a**). This finding suggests that a fraction of untreated sewage entered the groundwater system. The origin of the untreated sewage may have been from the surface or from a leaky sewage system. Torres-Hernández et al. (2020) used isotopic and chemical evidence to determine that nitrate pollution in groundwater from Monterrey was mainly derived from sewage leaks in urban areas. It is evident that organic and viral loads could have entered the groundwater system using the same pathway. The significant correlation between SARS-CoV-2 concentrations and sucralose at the 0.05 level is another remarkable confirmation of the contribution of raw wastewater to the groundwater and reaffirms possible leaching and infiltration of effluents from health care facilities, sewage, solid landfills, and drainage water as well as failing sewage pipes in the MMA (**Fig. 3c**).

From the three aquifer units used for water supply, the SA system (63%) was most affected, followed by the ZM aquifer (54%), and the BA well field (22%). Nevertheless, the viral loads observed in the wells of the first sampling campaign (29 October 2020 - 4 November 2020) were only partly reproduced one month later (26-30 November 2020), indicating a decrease in the viral load. This result demonstrates how dynamic the groundwater system is in relation to the presence of the coronavirus; the decline of the viral load in groundwater appeared to follow the decreasing trend in reported cases of infection during the month of November 2020 (**Fig. 2a**).

From the sampled municipalities in the MMA during the first campaign, Apodaca had the most positive samples at with 63% of the samples, followed by Monterrey (50%), and San Nicolas (50%). Coincidently, these are the most affected municipalities considering the officially reported daily cases of infection in **Fig. 2a**. Guadalupe was also among the most affected municipalities; however, it was represented by only one sampled well. Santiago, the southernmost municipality was the exception as it had a relatively lower number of cases of infection, but a high incidence of positive cases (63%). This scenario could indicate a different dynamic. Generally, the high number of positive samples in municipalities with highest number of COVID infections suggests that groundwater samples approximately mirror the infection situation at the municipality level.

### 4.4 Implications for public health

This study provides the first evidence that SARS-CoV-2 may enter groundwater through possible leaching events and infiltration of effluent from health care facilities, sewage, solid landfills and drainage water, as well as leakages from sewage pipes. Groundwater in the MMA is currently disinfected by gas chlorination removing pathogenic viruses and bacteria before entering the water supply system. Since coronaviruses are sensitive to oxidants such as chlorine (La Rosa et al., 2020b), it is important to continue strengthening and advancing the treatment processes of groundwater, especially in wells located in shallow aquifers and in places where sewage effluent from health care facilities, sewage, solid landfills and drainage water is not treated or treated inefficiently (Guerrero-Latorre et al. 2020) and is expected to infiltrate, or where sewage pipes could be leaky (Torres-Hernandez et al., 2020).

The concentrations of SARS-CoV-2 RNA in untreated wastewater from selected studies worldwide were in the range of not detected to 5600 GC/ml (**Table 5**). In our study, monitoring of the influent at the Dulces Nombres WWTP showed that between October 25, 2020, and December 13, 2020, 3 out of 8 samples (38%) were positive for SARS-CoV-2, and that the maximum load was 3535 GC/ml (**Fig. 3b**). This number is quite comparable to other studies of raw wastewater during outbreaks (Nemudryi et al., 2020; Randazzo et al. 2020; Torttier et al, 2020; Wurtzer et al., 2020; Medema et al., 2020; **Table 5**). This result shows that the concentration of SARS-CoV-2 in the surface water (<5.6 GC/ml) and groundwater (<38.3 GC/ml) in the MMA is approximately two to three orders of magnitude lower than that in raw wastewater. This means that the viral load could not be eradicated completely, as observed in Haramoto et al. (2020); however the result in this study is similar to that in Rimoldi et al. (2020).

The presence of SARS-CoV-2 genetic material in natural waters receiving treated or untreated wastewater effluents raises the important question of whether there is a risk of infection. Since urban water courses and dams are very popular places for recreation, there is concern about the risks of infection. The transmission potential of SARS-CoV-2 by ingestion is still controversial but potentially occurs (Amirian et al., 2020). Kumar et al. (2021) suggested a quantitative microbial risk assessment framework to estimate the potential risk from SARS-CoV-2 in natural water bodies through various water activities, based on the framework for SARS-CoV developed by Watanabe et al. (2010). The support for this approach is that there is no risk assessment model available for ingestion of water with SARS-CoV-2 and that both SARS-CoV and SARS-CoV-2 species have similar genetics and infection mechanisms. According to this approach, the chances of infection by a virus are calculated by a dose–response model, which describes relations of viral exposure dose and the probability of infection and can be calculated by an exponential model with the following equation:

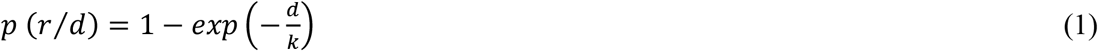

where *p* (*r*/*d*)is the chance of infection at the viral dose of *d, d* is dose of the virus (PFU, plaque-forming unit), and *k* is 4.2 × 10^2^ (PFU). The expected dose of the virus is estimated from the volume of water ingested and the viral concentration in the water. The median volume of water ingested per event is reported to be 6.0 ml when swimming and 2.0 ml when fishing (Dorevitch et al., 2011). Considering a viral load of 7.0 copies/ml in the rivers of MMA (**Fig. 3b**), the estimated chance of infection per event was derived from equation (1) as 1.0 × 10^-7^ for swimming and 3.4 × 10^-8^ for fishing.

These findings suggest a very low risk of transmission of SARS-CoV-2 during recreation in waters receiving treated wastewater from the MMA. However, the presence of detectable amounts of genetic material from SARS-CoV-2 in fresh water should not be ignored. There exist situations where the infection risk may increase considerably. For example, the occurrence of CSO events during COVID-19 outbreaks may cause a substantial increase in the infection risk of SARS-CoV-2 by exposure to receiving water bodies (Kumar et al., 2021). Another situation is that residual chlorine may not be maintained in sufficient concentration to control the virus. Consider a fictional case where raw wastewater from the Dulces Nombres WWTP with 3535 copies/ml is discharged into the riverbed without dilution as may occur during a drought period, then the chances of infection increase to 5.2 × 10^-5^ and 1.7 × 10^-5^ for swimming and fishing, respectively.

Under normal operating conditions, the infection risk in groundwater is minimal if the pumping wells are on a well seal, which protects it from surface contamination, and the disinfection system is working properly. However, in a leaky pumping well, the infiltration of human effluent spills combined with a failure in the disinfection system may considerably increase the infection risk. Assuming a sludge concentration of 10^5^ copies/ml infiltrates and is diluted 5 times in groundwater, then the immediate chance of infection from drinking a glass of untreated water is 0.15%. Therefore, an annual, preventative water-well maintenance inspection is important to avoid any risks of a COVID-19 infection through groundwater.

It is important to note that these values could be underestimated and have large uncertainty associated with them, because SARS-CoV-2 is potentially more infectious than SARS-CoV from which the model is derived (Kitajima et al., 2020). On the other hand, the proportion of viable RNA copies in the measured viral load was not known. SARS-CoV-2 RNA was found to be significantly more persistent than infectious SARS-CoV-2, indicating that the environmental detection of RNA alone does not substantiate the risk of infection (Bivins et al., 2020). Thus, this risk assessment model should be considered a preliminary estimation or base line of the associated health risks for SARS-CoV-2 in aquatic environments.

### 4.5 Limitations and future directions

This study shows the importance of monitoring programs to determine the fate of SARS-CoV-2 in the urban water cycle. To date, there is no evidence related to the fate of SARS-CoV-2 in the urban water cycle, and few datasets exist to confirm whether water or wastewater containing SARS-CoV-2 could be potentially infectious. Some studies have predicted a low risk of SARS-CoV-2 transmission via wastewater (Chin et al., 2020; Rimoldi et al., 2020), but this topic still deserves attention and further detailed examinations (Buonerba et al., 2021). It is necessary to monitor natural waters, especially in countries or areas that have limited capacities of wastewater treatment.

Future research should be oriented towards the development of a proper SARS-CoV-2 infection risk assessment model, considering the virus in its different variants. This model could be based on dose-response approaches developed for other pathogens (Watanabe et al. 2010; de Man et al., 2014) and use SARS-CoV-2 data sets yet to be developed from experiments.

Another area of opportunity is to study the SARS-CoV-2 removal efficiency of wastewater treatment processes including disinfection. One limitation of this study is a lack of understanding of how the removal efficiency of a WWTP contributes to the dilution of the viral load in the receiving river water. In general, there is still minimal knowledge about the removal of enveloped viruses in wastewater (Kumar et al., 2021).

The use of chemical and microbial markers for human wastewater could assist in not only evaluating the removal efficiency of wastewater treatment facilities but also understanding the routes and fates of SARS-CoV-2 in natural water systems. For biosafety purposes, surrogate viruses such as the murine hepatitis virus and phages were employed successfully due to their structural and morphological similarity to SARS-CoV-2 (e.g. Ahmed et al., 2020b). The combined use of selected markers could provide additional information about the dilution, decay, and inhibition factors of the new coronavirus in aquatic environments.

Studies performed to date show that there is a lack of standardized protocols for sampling, detecting and quantifying of SARS-CoV-2 in water and wastewater (**Table 5**). For example, in some studies grab samples were obtained, while in others 24-hour composite samples were collected. In this study we used a sample size (125 ml) and recognize that larger samples would be a more appropriate choice and that would have derived in a more representative finding. Also, the sampling duration was relatively short.

There were significant differences in not only sample collection but also sample storage and treatment and the use (or not) of genetic or chemical traces (i.e., chemical agents indicating human activity or viral tracers used for normalization purposes). This may lead to discrepancies in the results. Currently, RT-qPCR has been employed widely for detection of SARS-CoV-2 in water samples; however it is imperative to develop a standard sampling procedure for accurate extraction, isolation, detection and quantification of the virus. The N gene (N1&N2) is the most abundant transcript of SARS-CoV-2 and is therefore a good target for the detection of the virus in samples (Babiker et al., 2020; Chakraborty et al., 2021). Inter- and intralaboratory comparisons such as those employed by Chick et al. (2021) may lead to global standardization.

## 5. Conclusions

This study evaluated the presence of SARS-CoV-2 RNA in different freshwater areas of a metropolitan area and the implications for the environment and public health. As such, this study represents a contribution to the ongoing discussion on the potential routes and fate of SARS-CoV-2 in freshwater environments receiving wastewater and water safety concerns.

This is the first study that detected and quantified SARS-CoV-2 RNA in groundwater. Nearly half of the samples showed detectable genetic material. This result suggests that in a pump well, sewage from the surface or from a leaky sewage system entered the groundwater system. Moreover, the temporal and submetropolitan variations in the viral loads in groundwater mimic the reported trend in cases of infection in ZMM.

The share of detectable SARS-CoV-2 RNA in urban rivers (21%) and dams (12%) was lower than that in groundwater. The quantitative results show that the viral loads in these waters were three orders of magnitude lower than the maximum value measured in raw wastewater during the same time period. It is assumed that aliquots of nontreated sewage due to illegal discharges, local malfunctions of the sewage system and their increased relative contribution during the dry period may have been the factors. Again, there was a correlation between the temporal variation in the viral loads in the surface waters and the trend in the reported cases of infections. A preliminary risk assessment model suggests that, considering the viral loads found during this study in the receiving waters of Monterrey, the potential of infection was low for recreational activities (swimming, fishing, etc.). However, this situation should not be taken lightly because the occurrence of combined sewer overflow events and/or temporal failures of disinfection systems may cause substantial increases in infection risks.

This study shows that knowledge about the routes and fates of SARS-CoV-2 in the environment is still in the early stage and that datasets for water are scarce. In the short term, it is important to monitor especially natural water systems that receive untreated or poorly treated wastewaters. In the medium and long term, the COVID-19 pandemic represents an opportunity for the international community to accelerate the UN Sustainability Development Goal 6 (clean water and sanitation for all) by fostering financial and technical support to programs that increase the capacity of preventative water-well maintenance inspections and wastewater treatment, especially in less developed countries.

Future research and innovation efforts in this regard should be oriented towards: (i) the development of a proper SARS-CoV-2 infection risk assessment model for water and wastewater; (ii) an assessment of the removal efficiency of SARS-CoV-2 in wastewater treatment processes including disinfection; (iii) the combined use of chemical and microbiological markers for tracing the routes, decay and inhibition factors of SARS-CoV-2 in water; and (iv) the development of standardized protocols for sampling, detecting and quantifying SARS-CoV-2 in the environment.

## Data Availability

NA

## Acknowledgments

We would like to acknowledge the financial support received from Consejo Nacional de Ciencia y Tecnología (CONACyT) through Fondo Desarrollo Tecnológico e Innovación COVID-19 grant No. 312558. Logistical and technical support during the sampling campaign was provided by Servicio de Agua y Drenaje de Monterrey (SADM). A. Torres-Hernandez and Christian F. Narváez-Montoya assisted in the field work.

## Supplementary Material

**Table S1:**
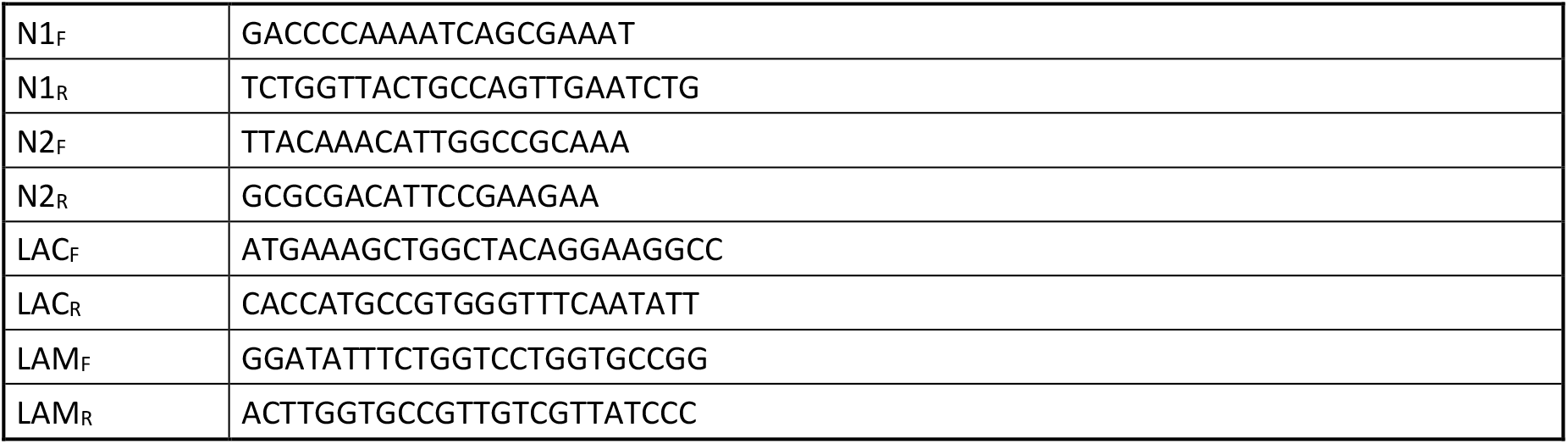
Sequences of the primer sets used for the detection of SARS-CoV-2 (N1 and N2) and E. coli (LAC and LAM) in residual and surface waters. The sequences of the forward and reverse primers are presented.

**Table S2:** Analytical conditions for the determination of sucralose

|  |  |
| --- | --- |
| HPLC system: | HPLC 1290 (Agilent Technologies) |
| Sample volume: | 50 mL |
| Sample pH: | 3 (adjusted with hydrochloric acid) |
| SPE material: | Strata X (200 mg / 6 mL) |
| Elution solvent: | 3 x 3 mL methanol |
| Reconstitution solvent: | 500 µL MilliQ water:methanol (80:20, v:v) |
| Injection volume: | 15 µL |
| Separation column: | ZORBAX Eclipse XDB-C8, 150 mm x 2.1 mm, 3.5 µm (Agilent Technologies) |
| Column temperature: | 40 °C |
| Eluents: | A: MilliQ water |
|  | B: Methanol |
| Additive to the eluent: | 10 mM ammonium acetate |
| Elution programme: | 0 min: 90% A 10% B |
|  | 2 min: 90% A 10% B |
|  | 6 min: 25% A 75% B |
|  | 11 min: 25% A 75% B |
|  | 12 min: 90% A 10% B |
| Flow rate: | 0.30 mL/min |
| MS-MS system: | Triple Quad 5500 (AB Sciex) |
| Interface: | Elektrospray (ESI) |
| Ionisation mode: | negative |
| Detection mode: | MRM (multi reaction monitoring) |
| Mass transitions: | Sucralose: 394.9 > 358.9 and 394.9 > 361.1 |

## References

Ahmed, W., Angel, N., Edson, J., Bibby, K., Bivins, A., O’Brien, J. W., … & Mueller, J. F. (2020a). First confirmed detection of SARS-CoV-2 in untreated wastewater in Australia: a proof of concept for the wastewater surveillance of COVID-19 in the community. Science of the Total Environment, 728, 138764.

Ahmed, W., Bertsch, P. M., Bibby, K., Haramoto, E., Hewitt, J., Huygens, F., … & Bivins, A. (2020b). Decay of SARS-CoV-2 and surrogate murine hepatitis virus RNA in untreated wastewater to inform application in wastewater-based epidemiology. Environmental Research, 191, 110092.

Amirian, E. S. (2020). Potential fecal transmission of SARS-CoV-2: Current evidence and implications for public health. International Journal of Infectious Diseases.

Babiker, A., Myers, C.W., Hill, C.E., Guarner, J., 2020. SARS-CoV-2 Testing: Trials and Tribulations. Oxford University Press US, 706–708.

Balboa, S., Mauricio-Iglesias, M., Rodríguez, S., Martínez-Lamas, L., Vasallo, F. J., Regueiro, B., & Lema, J. M. (2020). The fate of SARS-CoV-2 in wastewater treatment plants points out the sludge line as a suitable spot for incidence monitoring. MedRxiv.

Bar-Or, I., Yaniv, K., Shagan, M., Ozer, E., Erster, O., Mendelson, E., … & Kushmaro, A. (2020). Regressing SARS-CoV-2 sewage measurements onto COVID-19 burden in the population: a proof-of-concept for quantitative environmental surveillance. MedRxiv.

Bej, A. K., Mahbubani, M. H., Miller, R., DiCesare, J. L., Haff, L., & Atlas, R. M. (1990). Multiplex PCR amplification and immobilized capture probes for detection of bacterial pathogens and indicators in water. Molecular and Cellular Probes, 4(5), 353–365.

Benotti, M. J., Trenholm, R. A., Vanderford, B. J., Holady, J. C., Stanford, B. D., & Snyder, S. A. (2009). Pharmaceuticals and endocrine disrupting compounds in US drinking water. Environmental science &technology, 43(3), 597–603.

Betancourt, W. Q., Schmitz, B. W., Innes, G. K., Prasek, S. M., Brown, K. M. P., Stark, E. R., … & Pepper, I. L. (2021). COVID-19 containment on a college campus via wastewater-based epidemiology, targeted clinical testing and an intervention. Science of The Total Environment, 146408.

Bivins, A., Greaves, J., Fischer, R., Yinda, K. C., Ahmed, W., Kitajima, M., … & Bibby, K. (2020). Persistence of SARS-CoV-2 in water and wastewater. Environmental Science &Technology Letters, 7(12), 937–942.

Buerge, I. J., Buser, H. R., Kahle, M., Muller, M. D., & Poiger, T. (2009). Ubiquitous occurrence of the artificial sweetener acesulfame in the aquatic environment: an ideal chemical marker of domestic wastewater in groundwater. Environmental science &technology, 43(12), 4381–4385.

Buonerba, A., Corpuz, M. V. A., Ballesteros, F., Choo, K. H., Hasan, S. W., Korshin, G. V., … & Naddeo, V. (2021). Coronavirus in Water Media: Analysis, Fate, Disinfection and Epidemiological Applications. Journal of Hazardous Materials, 125580.

Chakraborty, P., Pasupuleti, M., Shankar, M. J., Bharat, G. K., Krishnasamy, S., Dasgupta, S. C., … & Jones, K. C. (2021). First surveillance of SARS-CoV-2 and organic tracers in community wastewater during post lockdown in Chennai, South India: Methods, occurrence and concurrence. Science of The Total Environment, 146252.

Chan, J. F. W., Yuan, S., Kok, K. H., To, K. K. W., Chu, H., Yang, J., … & Yuen, K. Y. (2020). A familial cluster of pneumonia associated with the 2019 novel coronavirus indicating person-to-person transmission: a study of a family cluster. The Lancet, 395(10223), 514–523.

Chik, A. H., Glier, M. B., Servos, M., Mangat, C. S., Pang, X. L., Qiu, Y., … &CoV-2 Inter-Laboratory Consortium. (2021). Comparison of approaches to quantify SARS-CoV-2 in wastewater using RT-qPCR: Results and implications from a collaborative inter-laboratory study in Canada. Journal of Environmental Sciences, 107, 218–229.

Chin, A., Chu, J., Perera, M., Hui, K., Yen, H. L., Chan, M., … & Poon, L. (2020). Stability of SARS-CoV-2 in different environmental conditions. MedRxiv.

CONACyT - Consejo Nacional de Ciencia y Tecnologia. (2021). Tablero de datos Coronavirus. Retrieved from: https://datos.covid-19.conacyt.mx/

De Man, H., Bouwknegt, M., van Heijnsbergen, E. J. T. M., Leenen, E. J. T. M., Van Knapen, F., &de Roda Husman M. (2014). Health risk assessment for splash parks that use rainwater as source water. Water Research, 54, 254–261.

Dorevitch, S., Panthi, S., Huang, Y., Li, H., Michalek, A. M., Pratap, P., … & Li, A. (2011). Water ingestion during water recreation. Water research, 45(5), 2020–2028.

Gasser, G., Rona, M., Voloshenko, A., Shelkov, R., Tal, N., Pankratov, I., … & Lev, O. (2010). Quantitative evaluation of tracers for quantification of wastewater contamination of potable water sources. Environmental science &technology, 44(10), 3919–3925.

González-González, E., Trujillo-de Santiago, G., Lara-Mayorga, I. M., Martinez-Chapa, S. O., & Alvarez, M. M. (2020). Portable and accurate diagnostics for COVID-19: Combined use of the miniPCR thermocycler and a well-plate reader for SARS-CoV-2 virus detection. PloS One, 15(8), e0237418.

González-González, E., Lara-Mayorga, I. M., Rodríguez-Sánchez, I. P., Zhang, Y. S., Martínez-Chapa, S. O., Trujillo-de Santiago, G., & Alvarez, M. M. (2021). Colorimetric loop-mediated isothermal amplification (LAMP) for cost-effective and quantitative detection of SARS-CoV-2: the change in color in LAMP-based assays quantitatively correlates with viral copy number. Analytical Methods.

Green, H., Wilder, M., Middleton, F. A., Collins, M., Fenty, A., Gentile, K., … & Larsen, D. A. (2020). Quantification of SARS-CoV-2 and cross-assembly phage (crAssphage) from wastewater to monitor coronavirus transmission within communities. MedRxiv.

Guerrero-Latorre, L., Ballesteros, I., Villacrés-Granda, I., Granda, M. G., Freire-Paspuel, B., &Ríos-Touma, B. (2020). SARS-CoV-2 in river water: Implications in low sanitation countries. Science of the Total Environment, 743, 140832.

Haramoto, E., Malla, B., Thakali, O., & Kitajima, M. (2020). First environmental surveillance for the presence of SARS-CoV-2 RNA in wastewater and river water in Japan. Science of The Total Environment, 737, 140405.

Hata, A., Honda, R., Hara-Yamamura, H., & Meuchi, Y. (2020). Detection of SARS-CoV-2 in wastewater in Japan by multiple molecular assays-implication for wastewater-based epidemiology (WBE). MedRxiv.

INEGI - Instituto Nacional de Estadística y Geografía. (2021). Censo de población y vivienda 2020. Retrieved from: https://censo2020.mx/

Kang, M., Wei, J., Yuan, J., Guo, J., Zhang, Y., Hang, J., … & Zhong, N. (2020). Probable evidence of fecal aerosol transmission of SARS-CoV-2 in a high-rise building. Annals of Internal Medicine, 173(12), 974–980.

Kitajima, M., Ahmed, W., Bibby, K., Carducci, A., Gerba, C. P., Hamilton, K. A., … & Rose, J. B. (2020). SARS-CoV-2 in wastewater: State of the knowledge and research needs. Science of The Total Environment, 139076.

Kocamemi, B. A., Kurt, H., Hacioglu, S., Yarali, C., Saatci, A. M., & Pakdemirli, B. (2020). First data-set on SARS-CoV-2 detection for Istanbul wastewaters in Turkey. MedRxiv.

Kokotou, M. G., Asimakopoulos, A. G., & Thomaidis, N. S. (2012). Artificial sweeteners as emerging pollutants in the environment: analytical methodologies and environmental impact. Analytical Methods, 4(10), 3057–3070.

Kumar, M., Patel, A. K., Shah, A. V., Raval, J., Rajpara, N., Joshi, M., & Joshi, C. G. (2020). First proof of the capability of wastewater surveillance for COVID-19 in India through detection of genetic material of SARS-CoV-2. Science of The Total Environment, 746, 141326.

Kumar, M., Alamin, M., Kuroda, K., Dhangar, K., Hata, A., Yamaguchi, H., &Honda, R. (2021). Potential discharge, attenuation and exposure risk of SARS-CoV-2 in natural water bodies receiving treated wastewater. npj Clean Water, 4(1), 1–11.

Langone, M., Petta, L., Cellamare, C. M., Ferraris, M., Guzzinati, R., Mattioli, D., & Sabia, G. (2021). SARS-CoV-2 in water services: presence and impacts. Environmental Pollution, 115806.

La Rosa, G., Iaconelli, M., Mancini, P., Ferraro, G. B., Veneri, C., Bonadonna, L., … & Suffredini, E. (2020a). First detection of SARS-CoV-2 in untreated wastewaters in Italy. Science of the Total Environment, 736, 139652.

La Rosa, G., Bonadonna, L., Lucentini, L., Kenmoe, S., & Suffredini, E. (2020b). Coronavirus in water environments: Occurrence, persistence and concentration methods-A scoping review. Water research, 115899.

Li, Q., Guan, X., Wu, P., Wang, X., Zhou, L., Tong, Y., … & Feng, Z. (2020). Early transmission dynamics in Wuhan, China, of novel coronavirus–infected pneumonia. New England Journal of Medicine.

Liu, J., Liao, X., Qian, S., Yuan, J., Wang, F., Liu, Y., … &Zhang, Z. (2020). Community transmission of severe acute respiratory syndrome coronavirus 2, Shenzhen, China, 2020. Emerging Infectious Diseases, 26(6), 1320.

Martinez, M. A. R., & Werner, J. (1997). Research into the Quaternary sediments and climatic variations in NE Mexico. Quaternary International, 43, 145–151.

Medema, G., Heijnen, L., Elsinga, G., Italiaander, R., & Brouwer, A. (2020). Presence of SARS-Coronavirus-2 RNA in sewage and correlation with reported COVID-19 prevalence in the early stage of the epidemic in the Netherlands. Environmental Science &Technology Letters, 7(7), 511–516.

Mlejnkova, H., Sovova, K., Vasickova, P., Ocenaskova, V., Jasikova, L., & Juranova, E. (2020). Preliminary study of Sars-Cov-2 occurrence in wastewater in the Czech Republic. International Journal of Environmental Research and Public Health, 17(15), 5508.

Mo, X. T., Zhou, Y. P., Lei, H., & Deng, L. (2002). Microbalance-DNA probe method for the detection of specific bacteria in water. Enzyme and Microbial Technology, 30(5), 583–589.

Mosley, L. M. (2015). Drought impacts on the water quality of freshwater systems; review and integration.Earth-Science Reviews, 140, 203–214.

Nalla, A. K., Casto, A. M., Huang, M. L. W., Perchetti, G. A., Sampoleo, R., Shrestha, L., … & Greninger, A. L. (2020). Comparative performance of SARS-CoV-2 detection assays using seven different primer-probe sets and one assay kit. Journal of Clinical Microbiology, 58(6).

Nemudryi, A., Nemudraia, A., Wiegand, T., Surya, K., Buyukyoruk, M., Cicha, C., … & Wiedenheft, B. (2020). Temporal detection and phylogenetic assessment of SARS-CoV-2 in municipal wastewater. Cell Reports Medicine, 1(6), 100098.

Oppenheimer, J., Eaton, A., Badruzzaman, M., Haghani, A. W., & Jacangelo, J. G. (2011). Occurrence and suitability of sucralose as an indicator compound of wastewater loading to surface waters in urbanized regions. Water research, 45(13), 4019–4027.

Peccia, J., Zulli, A., Brackney, D. E., Grubaugh, N. D., Kaplan, E. H., Casanovas-Massana, A., … & Omer, S. B. (2020). Measurement of SARS-CoV-2 RNA in wastewater tracks community infection dynamics. Nature Biotechnology, 38(10), 1164–1167.

Randazzo, W., Truchado, P., Cuevas-Ferrando, E., Simón, P., Allende, A., &Sánchez, G. (2020). SARS-CoV-2 RNA in wastewater anticipated COVID-19 occurrence in a low prevalence area. Water Research, 181, 115942.

Reza, Z. M., Mohammad, A., Salomeh, K., Reza, A. G., Hossein, S., Maryam, S., … & Saeed, F. (2014). Rapid detection of coliforms in drinking water of Arak city using multiplex PCR method in comparison with the standard method of culture (Most Probably Number). Asian Pacific Journal of Tropical Biomedicine, 4(5),404–409.

Rimoldi, S. G., Stefani, F., Gigantiello, A., Polesello, S., Comandatore, F., Mileto, D., … & Salerno, F. (2020). Presence and infectivity of SARS-CoV-2 virus in wastewaters and rivers. Science of the Total Environment, 744, 140911.

SADM - Servicios de Agua y Drenaje de Monterrey. (2020). Personal communication, 21 August 2020

SADM - Servicios de Agua y Drenaje de Monterrey. (2021). Personal communication, 28 January 2021

Sharif, S., Ikram, A., Khurshid, A., Salman, M., Mehmood, N., Arshad, Y., … & Ali, N. (2020). Detection of SARS-Coronavirus-2 in wastewater, using the existing environmental surveillance network: An epidemiological gateway to an early warning for COVID-19 in communities. MedRxiv.

Sherchan, S. P., Shahin, S., Ward, L. M., Tandukar, S., Aw, T. G., Schmitz, B., … & Kitajima, M. (2020). First detection of SARS-CoV-2 RNA in wastewater in North America: a study in Louisiana, USA. Science of The Total Environment, 743, 140621.

Sims, N., & Kasprzyk-Hordern, B. (2020). Future perspectives of wastewater-based epidemiology: monitoring infectious disease spread and resistance to the community level. Environment International, 139, 105689.

SMN - Servicio Meteorológico Nacional, Comisión Nacional del Agua (2020). Personal communication, 2021 January 28

Soh, L., Connors, K. A., Brooks, B. W., & Zimmerman, J. (2011). Fate of sucralose through environmental and water treatment processes and impact on plant indicator species. Environmental Science &Technology, 45(4), 1363–1369.

Torres-Martínez, J. A., Mora, A., Knappett, P. S., Ornelas-Soto, N., & Mahlknecht, J. (2020). Tracking nitrate and sulfate sources in groundwater of an urbanized valley using a multi-tracer approach combined with a Bayesian isotope mixing model. Water Research, 182, 115962.

Trottier, J., Darques, R., Mouheb, N. A., Partiot, E., Bakhache, W., Deffieu, M. S., & Gaudin, R. (2020). Post-lockdown detection of SARS-CoV-2 RNA in the wastewater of Montpellier, France. One Health, 10, 100157.

Voss, S., Newman, E., & Miller-Schulze, J. P. (2019). Quantification of sucralose in groundwater well drinking water by silylation derivatization and gas chromatography-mass spectrometry. Analytical Methods, 11(21), 2790–2799.

Wang, X., Zheng, J., Guo, L., Yao, H., Wang, L., Xia, X., & Zhang, W. (2020). Fecal viral shedding in COVID-19 patients: clinical significance, viral load dynamics and survival analysis. Virus Research, 289, 198147.

Watanabe, T., Bartrand, T. A., Weir, M. H., Omura, T., & Haas, C. N. (2010). Development of a dose‐response model for SARS coronavirus. Risk Analysis: An International Journal, 30(7), 1129–1138.

Westhaus, S., Weber, F. A., Schiwy, S., Linnemann, V., Brinkmann, M., Widera, M., … & Ciesek, S. (2021). Detection of SARS-CoV-2 in raw and treated wastewater in Germany–suitability for COVID-19 surveillance and potential transmission risks. Science of The Total Environment, 751, 141750.

WHO - World Health Organization (2020b). Modes of transmission of virus causing COVID-19: implications for IPC precaution recommendations: scientific brief, 29 March 2020 (WHO/2019-nCoV/Sci_Brief/Transmission_modes/2020.2). World Health Organization.

Wu, Y., Guo, C., Tang, L., Hong, Z., Zhou, J., Dong, X., … & Huang, X. (2020a). Prolonged presence of SARS-CoV-2 viral RNA in faecal samples. The Lancet Gastroenterology &hepatology, 5(5), 434–435.

Wu, F., Zhang, J., Xiao, A., Gu, X., Lee, W. L., Armas, F., … & Alm, E. J. (2020b). SARS-CoV-2 titers in wastewater are higher than expected from clinically confirmed cases. Msystems, 5 (4).

Wurtzer, S., Marechal, V., Mouchel, J. M., & Moulin, L. (2020). Time course quantitative detection of SARS-CoV-2 in Parisian wastewaters correlates with COVID-19 confirmed cases. MedRxiv.

Xiao, Y., Huang, S., Yan, L., Wang, H., Wang, F., Zhou, T., … & He, M. (2020). Clinical characteristics of diarrhea in 90 cases with COVID-19: A descriptive study. International emergency nursing, 52, 100912.

